# Multiscale dynamics of COVID-19 and model-based recommendations for 105 countries

**DOI:** 10.1101/2020.06.05.20123547

**Authors:** Jithender J. Timothy, Vijaya Holla, Günther Meschke

## Abstract

We analyse the dynamics of COVID-19 using computational modelling at multiple scales. For large scale analysis, we propose a 2-scale lattice extension of the classical SIR-type compartmental model with spatial interactions called the Lattice-SIRQL model. Computational simulations show that global quantifiers are not completely representative of the actual dynamics of the disease especially when mitigation measures such as quarantine and lockdown are applied. Furthermore, using real data of confirmed COVID-19 cases, we calibrate the Lattice-SIRQL model for 105 countries. The calibrated model is used to make country specific recommendations for lockdown relaxation and lockdown continuation. Finally, using an agent-based model we analyse the influence of cluster level relaxation rate and lockdown duration on disease spreading.

## 1 Introduction

### 1.1 Background

The severe acute respiratory syndrome coronavirus 2 (SARS-CoV-2), first identified in December 2019 in Wuhan, China, has since spread throughout the world, resulting in an ongoing pandemic, the Coronavirus disease 2019 (COVID-19) as a consequence of which the world is facing a serious and acute public health emergency. As of 5 June 2020, more than 6.63 million confirmed COVID-19 cases have been reported, resulting in more than 391,000 deaths around the world. Currently, there are no vaccines nor specific antiviral treatments for COVID-19. As a vaccine against SARS-CoV-2 is not expected before 2021, the current strategy in managing this pandemic is to decrease and delay the epidemic peak, known as ‘flattening the curve’. The curve here refers to the evolution of the number of active infectious individuals in time. Flattening the curve reduces the load on the health care system and slows down the rate of new infections.

Currently, strategies ranging from contact tracing of suspected infectious individuals, quarantine of confirmed cases and implementing various drastic contact mitigation measures ranging from social distancing and restricting public gatherings to shut-downs, lockdowns and limits on various activities have been enforced. While these measure have been implemented with the aim to reduce the risk of healthy individuals being infected and to decrease the load on the health-care system, these measures do impinge on civil liberties. Moreover, there has been a slowdown of unprecedented scale in economic activity (12). Due to COVID-19, the global economy is projected to contract by 3 % in 2020 with advanced economies losing upto 6% of their GDP (10). This translates to around 9 trillion dollars wiped out by COVID-19. The IMF has called this the worst decline since the Great Depression of the 1930s. Given the economic cost of the COVID-19 derived containment procedures, the efficiency of the current containment and lockdown measures are a subject of intense debate.

### 1.2 Modelling

Within this background our aim in this paper is to investigate the dynamics of COVID-19 using computational modelling. Mathematical models and computer simulations are useful experimental tools for testing scenarios and assessing hypothesis which however would not be practically and economically feasible by any other means. Moreover, COVID-19 is a new disease, and there is still a lot of uncertainty associated with its evolution.

Mathematical models for pandemics such as COVID-19 can help understand the course of the disease (5). Governments across the world are relying on projections to support decision-making during this pandemic. The most widely used model is the SIR model (14, 16). In its simplest avatar, the model describes the movement of individuals through three mutually exclusive stages of a disease: susceptible, infected and recovered at certain rate. For the ongoing COVID-19 pandemic, several SIR-type models have been used not only to predict the rate of spread of the virus (2,17) but also modified to take into account additional factors such as public risk perception and governmental action (18), age-structured interactions considering physical distancing (20), multiple discrete stages of the disease (11) and compartments representing mitigation measures (19). In general the more complex the model, higher are the number of parameters that have to be identified. Moreover, COVID-19 corresponds to an actively ongoing phenomena that makes calibration of the parameters even more challenging. While the aforementioned models do not explicitly model the dynamics of a single individual but only simulate collective dynamics, several models that consider realistic individual dynamics such as person-person contact etc. are currently in use for predicting the evolution of the disease (6). The most famous of which is the Ferguson Model (8) whose projections forced the UK government to abandon its plan of pursuing herd-immunity in favour of lockdown restrictions (1, 3). In such models a high degree of realism is made possible by introducing interactions and mechanisms at the scale of individual-individual interactions. However in introducing such detail, the model also requires additional parameters that govern these mechanisms. For e.g. It is much more difficult to predict the actions of a single individual than that of groups. Moreover we would like to stress the fact that COVID-19 is an ongoing disease and this makes it quite challenging to make realistic quantitative projections using such models (15).

While effective-medium compartmental models simulate the dynamics of the whole population using SIR-type dynamics which is not completely representative of the dynamics of the disease because the spatial component is completely missing in these types of model, the individual-based models are restricted by the complexity of the model i.e. restricted by the number of individuals they can simulate, computational power and parameters for the model. Our approach combines the strengths of both these modelling strategies by using a multiscale strategy.

## 2 Lattice-SIRQL

The model is a simple 2-scale extended SIR-model that includes additional states such as quarantine, lockdown and spatial interactions. Let *N_T_* be the total population of a country. In our multiscale model, this population is assumed to be comprised of *c* population clusters (cities, states, towns) such that 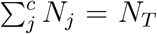. Where *N_j_* is the microscopic population of the *j^th^*cluster. At time *t* let 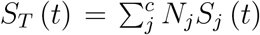 (t) be the total number of susceptibles in the population, 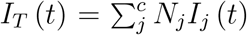 be the total number of infecteds, 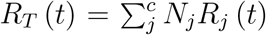 the total number of recovered, 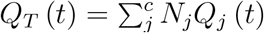 the total number of tested and quarantined confirmed cases and 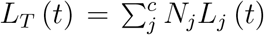 the total number of individuals under lockdown/containment. The quantities with subscripts *j* are cluster-specific normalized microscopic quantities. Given the microscopic quantities, the macroscopic quantities can be computed. These macroscopic quantities in turn satisfy the following balance relation (analogous to momentum or mass-balance in mechanics):

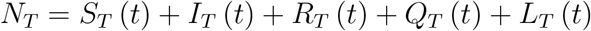

By specifying the balance equation as an initial condition (at the microscopic scale either directly or by down-scaling from the macroscopic scale), the dynamics of COVID-19 is then described by the time-dependent evolution of the 5 fields. The set of coupled differential equations (also balanced) that describe the evolution of the disease in a population with *c* clusters is then given by:

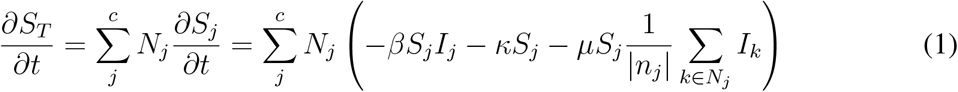

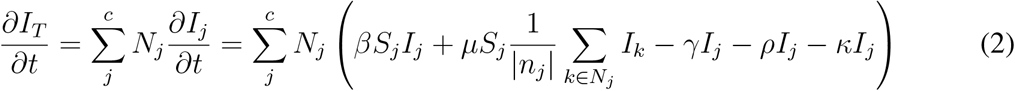

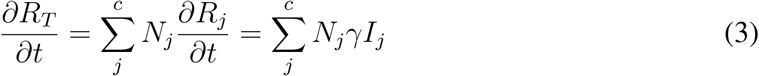

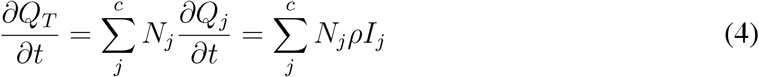

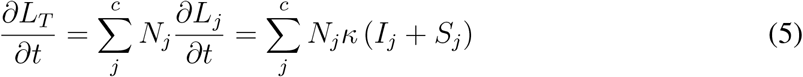

The first term on the right hand side of Eq.1 is the decrease in the susceptibles that is proportional to the product of the number of infectious individuals and the susceptibles with the proportionality constant being the transmission rate constant *β* and the second term is the decrease in susceptibles due to isolation and lockdown procedures with rate constant *κ* of individuals moving into lockdown and quarantine. The term 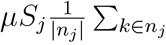 *I_k_* in Eq.1 is the increase in infections with rate constant *μ* due to interactions with neighbouring clusters. The set of all neighbouring clusters is denoted by *n_j_* and *|n_j_|* the total number of nearest neighbours. In Eq.2 the first and second terms on the right hand side is the proportional increase in the number of new infections (analogous to the decrease in the susceptibles in Eq.1). The third term is the decrease in the active infectious population due to recovery with rate constant *γ*. The fourth term is the decrease in the active infectious population due to testing and quarantine with the rate constant *ρ* and the last term is the decrease in the active infectious population due to self-isolation and government ordered lockdown. Finally, in order to solve the above expression, initial conditions have to be specified in terms of the number of initial infectious individuals. The inter-cluster interactions are computed using lattice-level convolution operations to compute nearest neighbour interactions in contrast to spatially discretized differential operators.

### 2.1 The reproduction number R

The main motivation for the widespread use of the SIR-type compartmental models is the ability of the model to characterize in simple terms a fundamental epidemiological quantity, i.e., the basic reproduction number (noted *R*_0_). This number is often misinterpreted and misunderstood. Firstly it is not a rate. Secondly it is only valid at the initial stage (the subscript 0 refers to the initial state) of disease subject to certain assumptions. In case of the classical SIR model (i.e assuming *μ* = 0, *ρ* = 0 and *κ* = 0 in Eq. 2 and dropping all subscripts and summations), the rate of change of active infections can be written as:

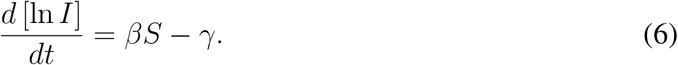

The exponential rate of change of *I* is proportional to the term *βS − γ*. If *βS > γ*, the infection grows exponentially and if *βS < γ*, the infection decreases exponentially. Thus we can mathematically represent these inequalities in terms of ratios with *R* = *βS/γ*. This quantity is indeed time dependent. i.e. depends on *S*. Assuming that initially the whole population is susceptible i.e. *S*_0_ = 1, *R*_0_ = *β/γ*. However, it must be noted that this particular definition is for a situation where the whole population is susceptible and no other mitigation procedures are in place. Moreover, it is also assumed that the transmissibility *β* is a constant. The reproduction number *R* is not something that is determined strictly by the virus (i.e. biological) as is portrayed often in public discourse but it is also determined by additional factors such as environmental, behavioural, and social characteristics. As lockdown and quarantine procedures start much later during the evolution of the disease, in this paper the definition of the reproduction number is assumed to be *R*_0_ = *β/γ*.

### 2.2 Flattening the Curve: Quarantine vs Lockdown

There are two ways in which the curve that corresponds to the number of active infectious members can be flattened. Either quarantine the infected members or lockdown the whole population. The latter option has serious economic consequences. In this subsection we investigate using a computational experiment the effect of quarantine vs lockdown on the evolution of the disease i.e. flattening the curve. We show that global averaged indicators of the disease do not reveal the complete picture because such indicators average out spatially localized concentrations of the disease that might potentially extend the lifetime of the disease.

1. Simulation Setup

a. The simulation consists of a population *N_T_* that is partitioned into *c* = 40000 clusters. The population per cluster is *N_j_* = *N_T_ /c* as we assume for simplicity a constant number of members per cluster. The number of initial infecteds is *I_T_* (*t* = 0) is assumed to be 0.0025% of *N_T_*. These infecteds are then further equally distributed randomly in a fraction *ϕ* = 0.02 of the *c* clusters (here, approximately 800 clusters). Thereafter we compute the normalized initial number of infecteds per cluster (the ones that belong to the group of 800 clusters) as *I_j_* = *I_T_ /* (ϕN_T_). Please note that this is the initial condition. We have omitted subscripts or additional notation to avoid cluttering. Finally the initial number of susceptibles is computed from the following expression assuming all other quantities are zero *S_j_* +*I_j_* +*R_j_* +*Q_j_* +*L_j_* = 1. Given the microscopic quantities, the macroscopic quantities are obtained by computing for e.g. 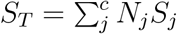. The time period of the simulation is 90 days.
2. The reference simulation

a. The basic reproduction number for the reference simulation is *R*_0_ = 3. The inter-cluster transmission rate constant *μ* is assumed to be 10% of the intra-cluster transmission rate constant *β* = 0.75. The time to recovery is assumed to be 7 days that correspond to *γ* = 1*/*7. The lockdown and quarantine rate constants are assumed to be zero (*κ* = 0, *ρ* = 0).
3. Quarantine only

a. We assume the same conditions as for the reference case but assuming a non-zero rate constant for quarantine with *ρ* = 0.25.
4. Lockdown only

a. We assume the same conditions as for the reference case but assuming a non-zero rate constant for lockdown with *κ* = 0.025. The rationale for choosing these values will be discussed later.

The top-most row in Fig. 2 shows the global macroscopic evolution of the three cases i.e. without mitigation measures, with quarantine and with lockdown. The 2D images correspond to the distribution and fraction of active infections (members that can potentially infect susceptibles) per cluster in the population. We have deliberately chosen a smaller value for *κ* = 0.025 because larger values lead to a dramatic decrease in infecteds in the simulation. This cannot be visualized and compared with the reference simulation using the same scale. Moreover, from a realistic point of view it would be much easier (with respect to logistics) to quarantine the smaller infected population than lockdown the whole population.

**Figure 1:**
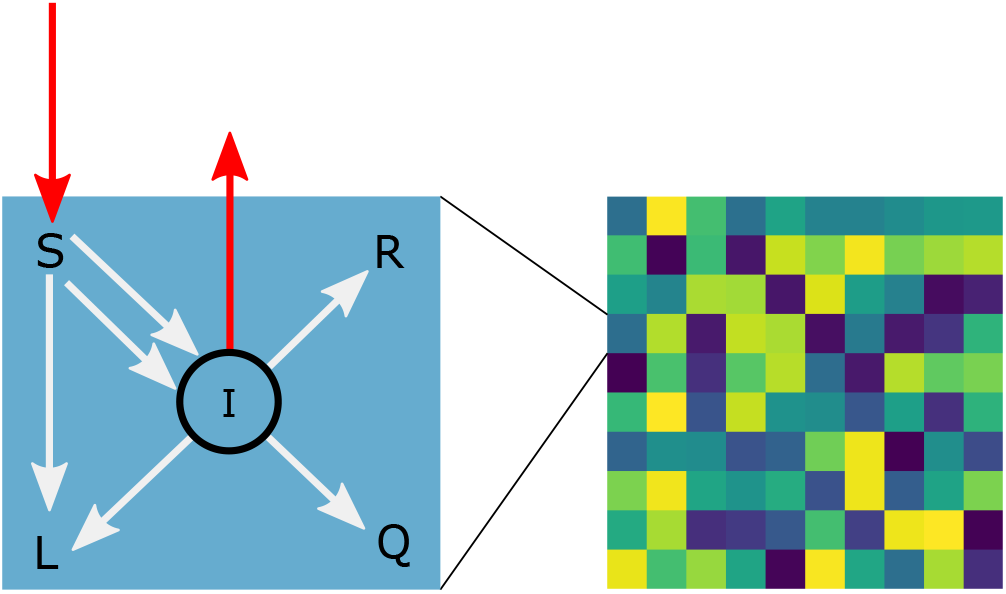
Schematic of the Lattice-SIRQL model. Left: Cluster level compartmental model. Right: Lattice of clusters that interact using nearest neighbour interactions.

**Figure 2:**
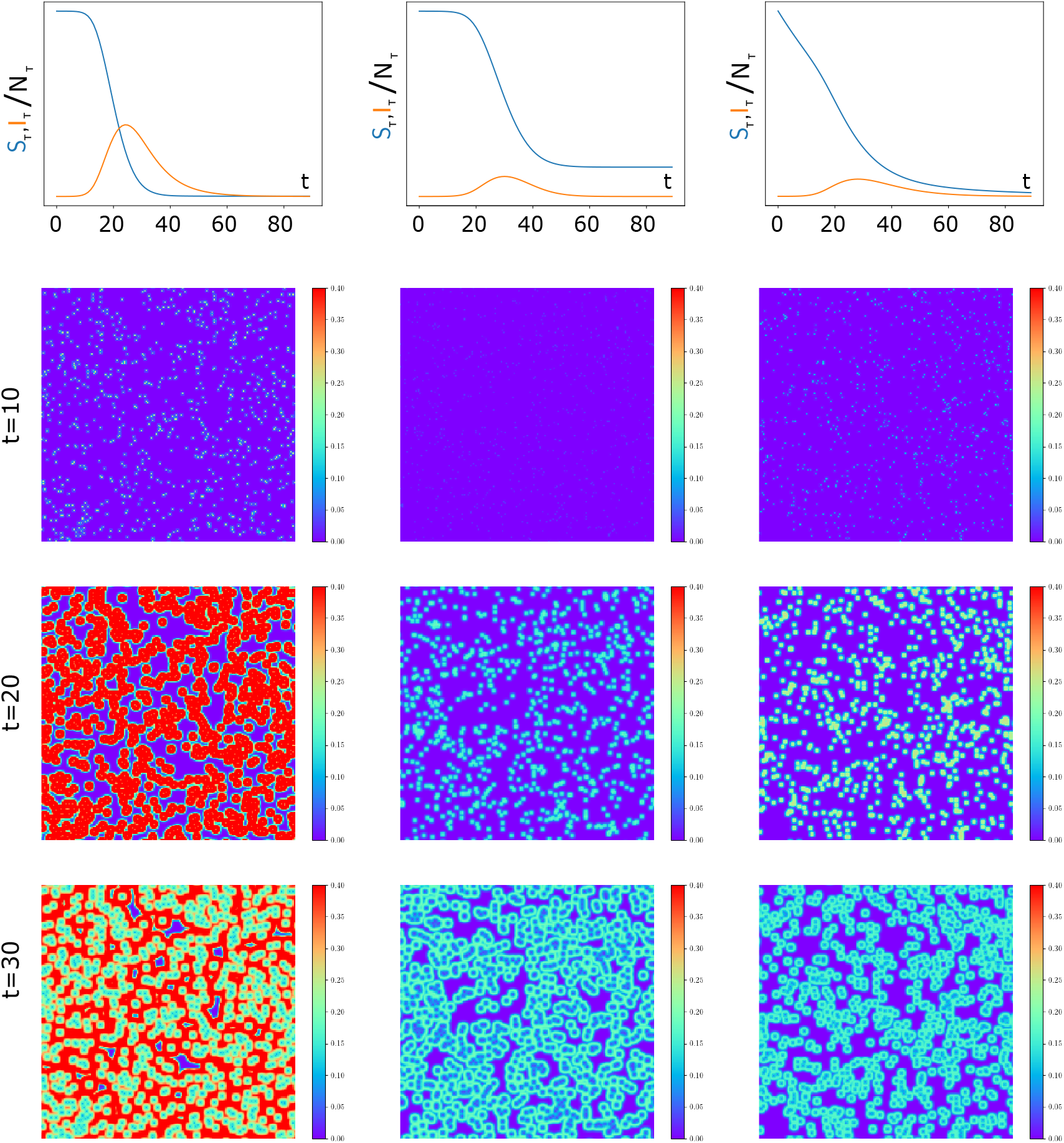
Simulation and comparison of the evolution of a pandemic without mitigation (left column), with quarantine only (center column) and with lockdown (right column) using the Lattice-SIRQL model. The top most curve is the global evolution of the susceptibles and the active infecteds in the population while the 2D images are spatially resolved simulation of the fraction of active infections per cluster after 10, 20 and 30 days.

It can be seen that for the case of quarantine and lockdown, eventhough showing approximately close peaks with regards to total number of active infecteds, the manner in which the curve corresponding to the number of active infections is flattened are completely different. While quarantine removes the infected population from mixing with the normal population, lockdown reduces interactions using social-distancing like measures by removing both infecteds and susceptibles from active mixing due to the coupling terms that are included in Eq. 1 and Eq.2 but not in Eq.4 and Eq.5. The curves corresponding to the number of susceptible members are also completely different for both cases. After 10 days, while for the quarantine case, the active infectious members have been removed, these infectious members are still active in the case of lockdown due to the smaller rate of removal and thus is reflected in the corresponding images. However, irrespective of this slow rate of removal in case of lockdown only, it is equally effective in reducing the total number of active infectious cases at the global level.

**Figure 3:**
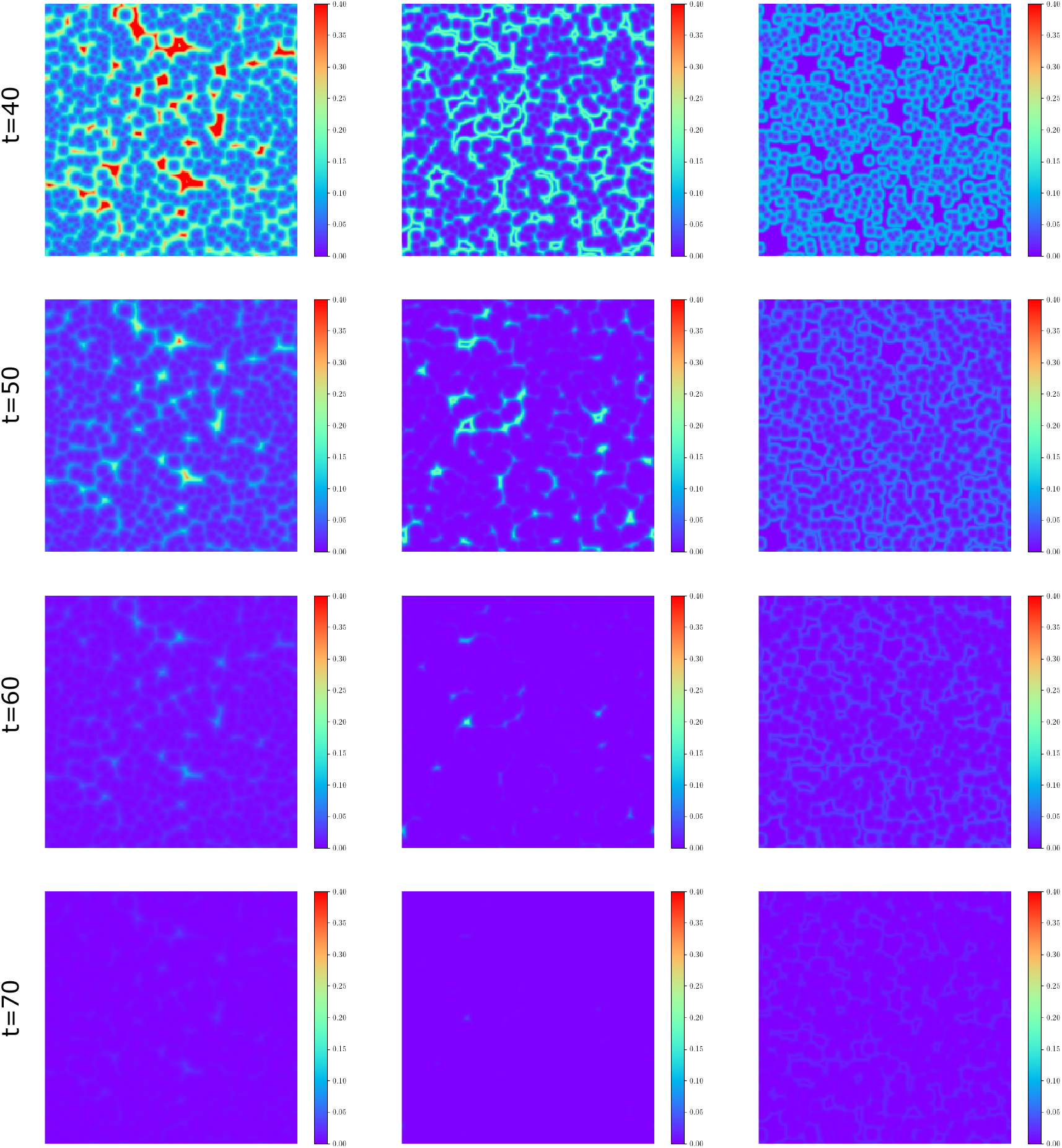
Simulation and comparison of the evolution of a pandemic without mitigation (left column), with quarantine only (center column) and with lockdown (right column) using the Lattice-SIRQL model. The 2D images are spatially resolved simulation of the fraction of active infections per cluster after 40, 50, 60 and 70 days.

After 40 days, while the case of quarantine shows hot-spots, lockdown has a lower concentration of infectious members. These hotspots in case of quarantine only remain until 60 days while lockdown has eliminated such hot-spots already after 30 days. Thus, just quarantine can significantly increase the risk of a second wave. Moreover, the simulation shows that at the global scale, quarantine measures and lockdown measures can show similar behaviour (the top most curves in Fig. 2) but their microscopic dynamics are different. This might not otherwise be evident by looking at spatially homogenized indicators of the pandemic. We at this point can only make a qualitative statement about the relevance of microscopic dynamics because the rate constants associated with the lockdown and quarantine are very difficult to quantify for a real case. However, in the next part of the paper we attempt to calibrate these parameters for over 105 countries.

## 3 Country-Specific Simulation, Calibration and model-based Recommendation

The confirmed number of infectious cases reported daily is limited by the testing capacity and does not reflect the true number of infectious individuals. Moreover, once a case is confirmed, it is also quarantined. Hence, the number of confirmed cases that are reported are assumed to be represented in the proposed model by *Q_T_*. We assume *I_T_* to be a ‘hidden’ variable.

In order to use the aforementioned model for a certain country: the rate constants associated with transmission (β, μ), recovery (γ), quarantine (ρ) and lockdown (κ) are required. Finally, initial conditions are required. Due to the varying demographic construction of each country and the level of isolation procedures and the testing capacity, the parameters associated with the rate constants should not be expected to be the same. We make the following assumptions in the model

1. The reported country specific data that we use to calibrate the model is assumed to correspond to infected individuals in the population who are symptomatic and quarantined *Q_T_*. These individuals are isolated from the general population and would not further infect any susceptible individuals.
2. The rate of recovery of an infected individual is also assumed to be fixed for all countries.

Given a) the fraction of infected quarantined cases, and b) the rate of recovery we compute the rate constants associated with transmission (intra- and inter-cluster), quarantine and lockdown for COVID-19. Table 1 lists the quantities that are fixed in the procedure to compute the rate constants.

**Table 1:** List of Fixed Parameters

| Parameter | value | source |
| --- | --- | --- |
| $\gamma$ | 1/7 | - |
| $N_T$ | country specific | Ref. (21) |
| $Q$ | country specific | Ref. (9) |

Still to be specified are the initial conditions. We assume the whole population of the country to be susceptible and the initial number of infected individuals to be proportional to the number of confirmed cases. Having specified these quantities, the model is complete.

### 3.1 Country-specific parameters

The country-specific model parameters are obtained by minimizing the error (in the least-squared sense) between the model predictions and the field data. Given the country specific time-series data corresponding to the number of confirmed cases 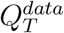 for a certain time period and the quantity *Q_T_* obtained by solving Eq. (1–5) for the same time period, the minimization problem is defined as:

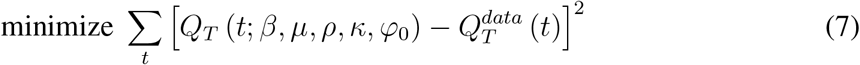

subject to the constraints *γ ≤ β ≤* 7*γ*, 0 *≤ μ ≤ β*, 0 *≤ ρ ≤* 1, 0 *≤ κ ≤* 1, 1 *≤ ϕ*_0_ *≤* 10^3^. The second constraint 0 *≤ μ ≤ β* is normalized and written as 0 *≤ μ^0^≤* 1, where *μ* = *μ^0^β*. We use the dual annealing optimization algorithm to solve this minimization problem. The dual annealing algorithm is a variation of the classical simulated annealing algorithm that is used for non-convex problems to find the global minimum of a function using metropolis-type updates. The principle of simulated annealing is motivated from statistical physics where a point in the parameter space is proposed with a certain distribution and this is accepted if it minimizes the residual and if not still accepted with a certain probability that is proportional to a certain ‘temperature’ that decreases with time. As temperature goes to zero, the algorithm behaves like the steepest descent algorithm. See (22) for further details. The maximum number of functional evaluations i.e number of times Eq. (1–5) is solved for the given time period per country is 100000. We solve the optimization problem on fully-parallel 80 multi threaded CPUs using the python multiprocessing module.

Figures 4, 5 and 6 show the reported data of confirmed COVID-19 cases in color along with the calibrated simulation curves. Countries that have not reported atleast 1000 cases have not been included in the analysis. The parameters identified by the minimization procedure is provided in Table 2. It must be noted that this is not a purely curve-fitting procedure but a procedure to fit reported field data to a physical model with a certain microscopic dynamics. It must be noted that the quality of the identified parameters is completely governed by the quality and reliability of the data. While the model predictions are predominantly in good agreement with reported data, there are some cases where the model predictions do not strictly conform to the trend of the reported data especially for Iceland inFig. 5. In general, the calibrated curves show that for the range of the chosen parameters, the model is able to capture all varieties of non-linearity associated with the reported data at the global level.

**Table 2:**
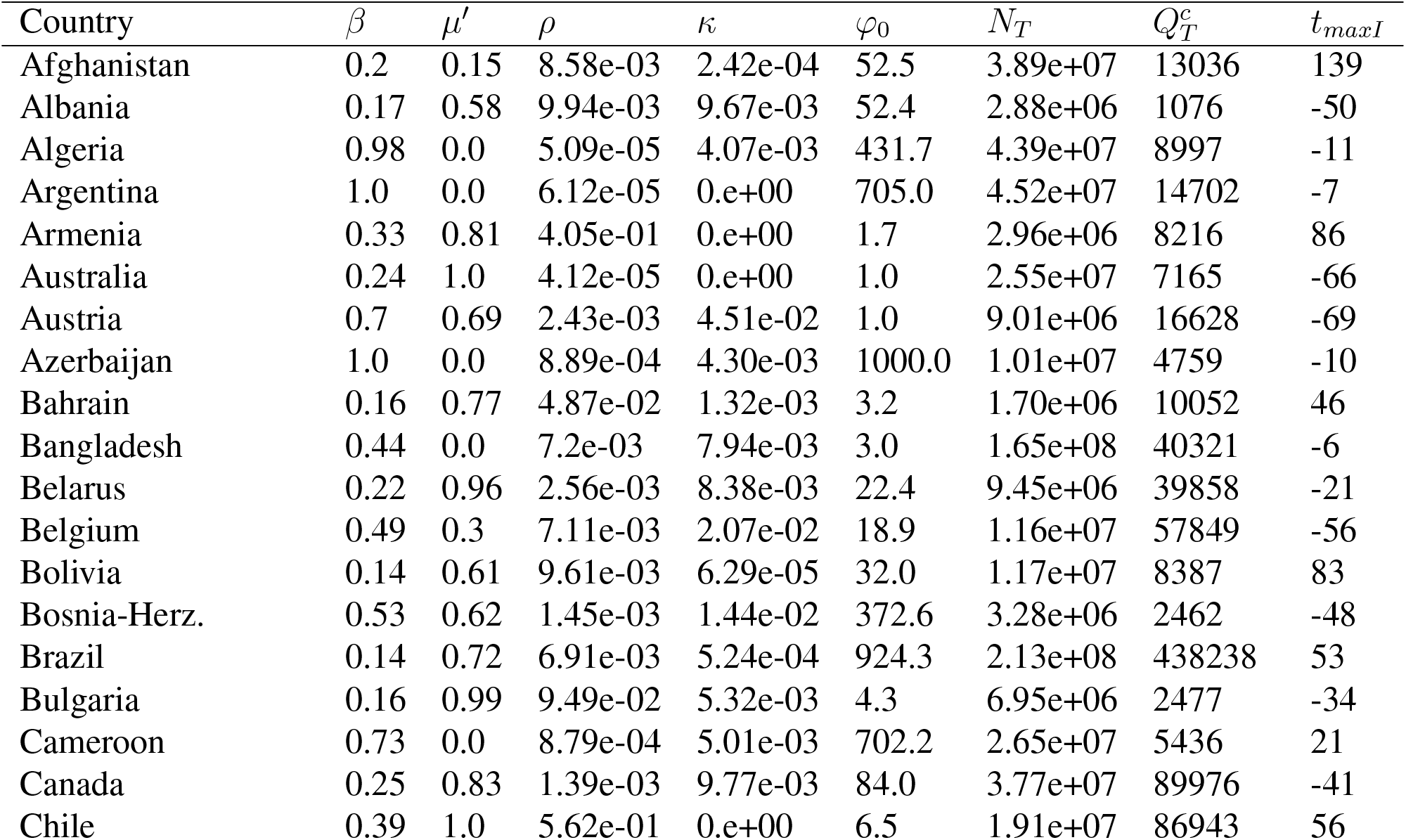
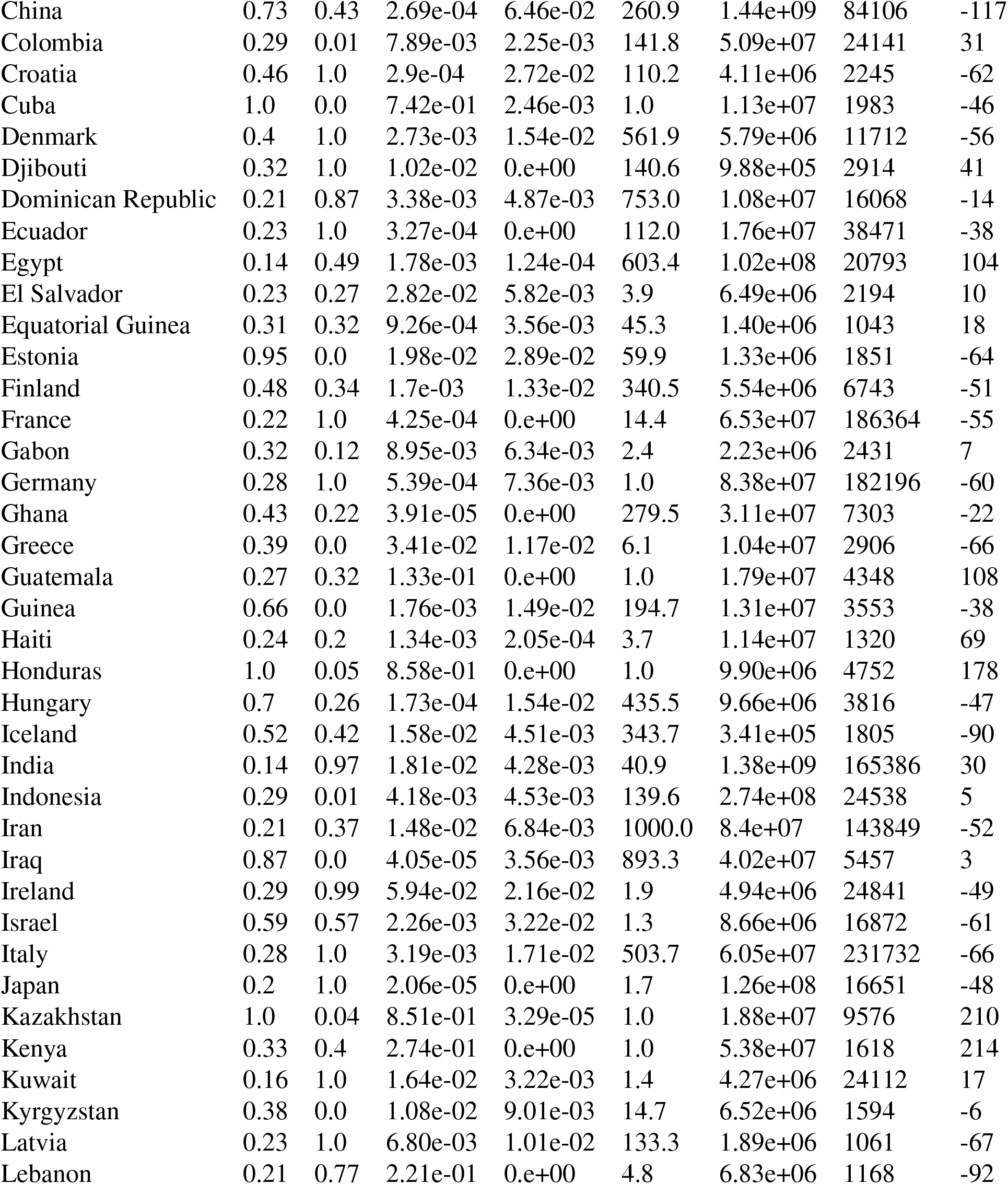
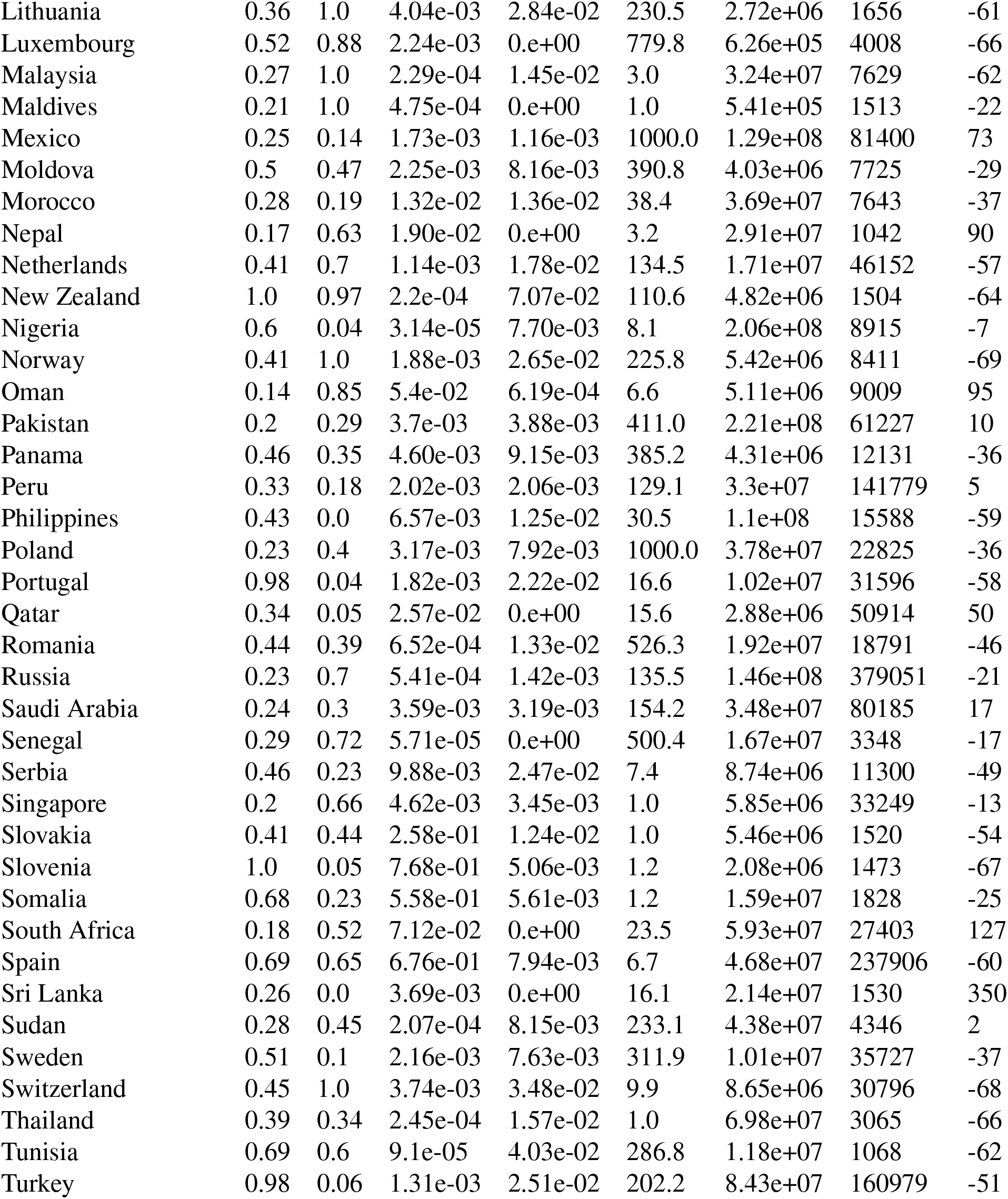
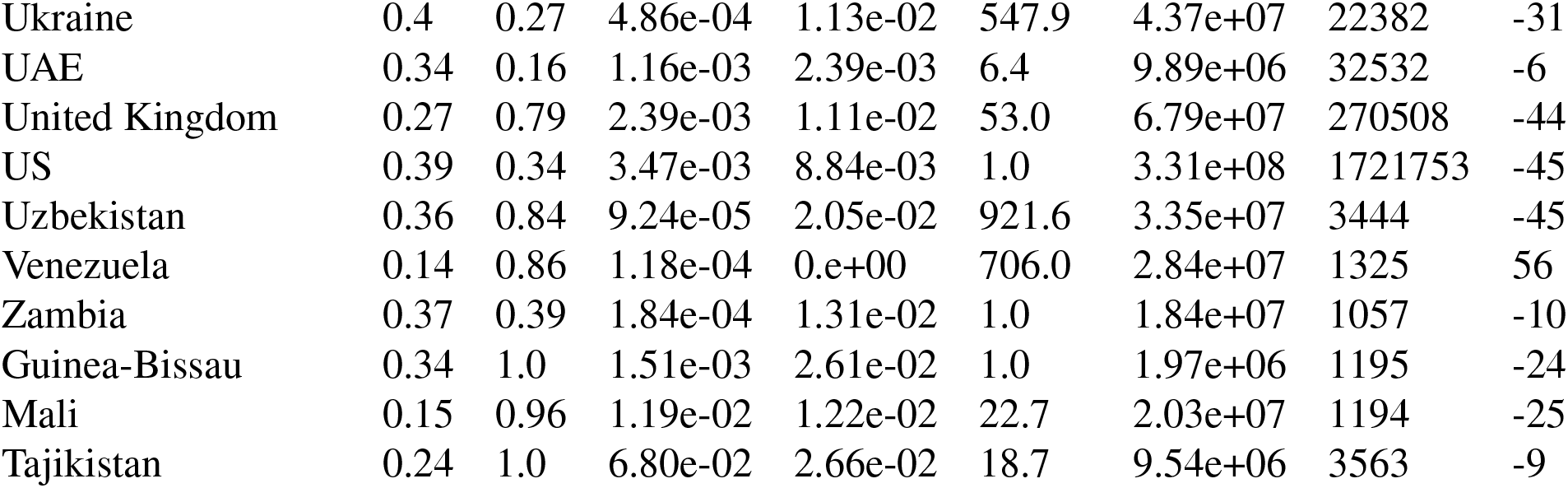
Country-specific model parameters

| Country | $\beta$ | $\mu'$ | $\rho$ | $\kappa$ | $\varphi_0$ | $N_T$ | $Q_T^c$ | $t_{maxI}$ |
| --- | --- | --- | --- | --- | --- | --- | --- | --- |
| Afghanistan | 0.2 | 0.15 | 8.58e-03 | 2.42e-04 | 52.5 | 3.89e+07 | 13036 | 139 |
| Albania | 0.17 | 0.58 | 9.94e-03 | 9.67e-03 | 52.4 | 2.88e+06 | 1076 | -50 |
| Algeria | 0.98 | 0.0 | 5.09e-05 | 4.07e-03 | 431.7 | 4.39e+07 | 8997 | -11 |
| Argentina | 1.0 | 0.0 | 6.12e-05 | 0.e+00 | 705.0 | 4.52e+07 | 14702 | -7 |
| Armenia | 0.33 | 0.81 | 4.05e-01 | 0.e+00 | 1.7 | 2.96e+06 | 8216 | 86 |
| Australia | 0.24 | 1.0 | 4.12e-05 | 0.e+00 | 1.0 | 2.55e+07 | 7165 | -66 |
| Austria | 0.7 | 0.69 | 2.43e-03 | 4.51e-02 | 1.0 | 9.01e+06 | 16628 | -69 |
| Azerbaijan | 1.0 | 0.0 | 8.89e-04 | 4.30e-03 | 1000.0 | 1.01e+07 | 4759 | -10 |
| Bahrain | 0.16 | 0.77 | 4.87e-02 | 1.32e-03 | 3.2 | 1.70e+06 | 10052 | 46 |
| Bangladesh | 0.44 | 0.0 | 7.2e-03 | 7.94e-03 | 3.0 | 1.65e+08 | 40321 | -6 |
| Belarus | 0.22 | 0.96 | 2.56e-03 | 8.38e-03 | 22.4 | 9.45e+06 | 39858 | -21 |
| Belgium | 0.49 | 0.3 | 7.11e-03 | 2.07e-02 | 18.9 | 1.16e+07 | 57849 | -56 |
| Bolivia | 0.14 | 0.61 | 9.61e-03 | 6.29e-05 | 32.0 | 1.17e+07 | 8387 | 83 |
| Bosnia-Herz. | 0.53 | 0.62 | 1.45e-03 | 1.44e-02 | 372.6 | 3.28e+06 | 2462 | -48 |
| Brazil | 0.14 | 0.72 | 6.91e-03 | 5.24e-04 | 924.3 | 2.13e+08 | 438238 | 53 |
| Bulgaria | 0.16 | 0.99 | 9.49e-02 | 5.32e-03 | 4.3 | 6.95e+06 | 2477 | -34 |
| Cameroon | 0.73 | 0.0 | 8.79e-04 | 5.01e-03 | 702.2 | 2.65e+07 | 5436 | 21 |
| Canada | 0.25 | 0.83 | 1.39e-03 | 9.77e-03 | 84.0 | 3.77e+07 | 89976 | -41 |
| Chile | 0.39 | 1.0 | 5.62e-01 | 0.e+00 | 6.5 | 1.91e+07 | 86943 | 56 |
| China | 0.73 | 0.43 | 2.69e-04 | 6.46e-02 | 260.9 | 1.44e+09 | 84106 | -117 |
| Colombia | 0.29 | 0.01 | 7.89e-03 | 2.25e-03 | 141.8 | 5.09e+07 | 24141 | 31 |
| Croatia | 0.46 | 1.0 | 2.9e-04 | 2.72e-02 | 110.2 | 4.11e+06 | 2245 | -62 |
| Cuba | 1.0 | 0.0 | 7.42e-01 | 2.46e-03 | 1.0 | 1.13e+07 | 1983 | -46 |
| Denmark | 0.4 | 1.0 | 2.73e-03 | 1.54e-02 | 561.9 | 5.79e+06 | 11712 | -56 |
| Djibouti | 0.32 | 1.0 | 1.02e-02 | 0.e+00 | 140.6 | 9.88e+05 | 2914 | 41 |
| Dominican Republic | 0.21 | 0.87 | 3.38e-03 | 4.87e-03 | 753.0 | 1.08e+07 | 16068 | -14 |
| Ecuador | 0.23 | 1.0 | 3.27e-04 | 0.e+00 | 112.0 | 1.76e+07 | 38471 | -38 |
| Egypt | 0.14 | 0.49 | 1.78e-03 | 1.24e-04 | 603.4 | 1.02e+08 | 20793 | 104 |
| El Salvador | 0.23 | 0.27 | 2.82e-02 | 5.82e-03 | 3.9 | 6.49e+06 | 2194 | 10 |
| Equatorial Guinea | 0.31 | 0.32 | 9.26e-04 | 3.56e-03 | 45.3 | 1.40e+06 | 1043 | 18 |
| Estonia | 0.95 | 0.0 | 1.98e-02 | 2.89e-02 | 59.9 | 1.33e+06 | 1851 | -64 |
| Finland | 0.48 | 0.34 | 1.7e-03 | 1.33e-02 | 340.5 | 5.54e+06 | 6743 | -51 |
| France | 0.22 | 1.0 | 4.25e-04 | 0.e+00 | 14.4 | 6.53e+07 | 186364 | -55 |
| Gabon | 0.32 | 0.12 | 8.95e-03 | 6.34e-03 | 2.4 | 2.23e+06 | 2431 | 7 |
| Germany | 0.28 | 1.0 | 5.39e-04 | 7.36e-03 | 1.0 | 8.38e+07 | 182196 | -60 |
| Ghana | 0.43 | 0.22 | 3.91e-05 | 0.e+00 | 279.5 | 3.11e+07 | 7303 | -22 |
| Greece | 0.39 | 0.0 | 3.41e-02 | 1.17e-02 | 6.1 | 1.04e+07 | 2906 | -66 |
| Guatemala | 0.27 | 0.32 | 1.33e-01 | 0.e+00 | 1.0 | 1.79e+07 | 4348 | 108 |
| Guinea | 0.66 | 0.0 | 1.76e-03 | 1.49e-02 | 194.7 | 1.31e+07 | 3553 | -38 |
| Haiti | 0.24 | 0.2 | 1.34e-03 | 2.05e-04 | 3.7 | 1.14e+07 | 1320 | 69 |
| Honduras | 1.0 | 0.05 | 8.58e-01 | 0.e+00 | 1.0 | 9.90e+06 | 4752 | 178 |
| Hungary | 0.7 | 0.26 | 1.73e-04 | 1.54e-02 | 435.5 | 9.66e+06 | 3816 | -47 |
| Iceland | 0.52 | 0.42 | 1.58e-02 | 4.51e-03 | 343.7 | 3.41e+05 | 1805 | -90 |
| India | 0.14 | 0.97 | 1.81e-02 | 4.28e-03 | 40.9 | 1.38e+09 | 165386 | 30 |
| Indonesia | 0.29 | 0.01 | 4.18e-03 | 4.53e-03 | 139.6 | 2.74e+08 | 24538 | 5 |
| Iran | 0.21 | 0.37 | 1.48e-02 | 6.84e-03 | 1000.0 | 8.4e+07 | 143849 | -52 |
| Iraq | 0.87 | 0.0 | 4.05e-05 | 3.56e-03 | 893.3 | 4.02e+07 | 5457 | 3 |
| Ireland | 0.29 | 0.99 | 5.94e-02 | 2.16e-02 | 1.9 | 4.94e+06 | 24841 | -49 |
| Israel | 0.59 | 0.57 | 2.26e-03 | 3.22e-02 | 1.3 | 8.66e+06 | 16872 | -61 |
| Italy | 0.28 | 1.0 | 3.19e-03 | 1.71e-02 | 503.7 | 6.05e+07 | 231732 | -66 |
| Japan | 0.2 | 1.0 | 2.06e-05 | 0.e+00 | 1.7 | 1.26e+08 | 16651 | -48 |
| Kazakhstan | 1.0 | 0.04 | 8.51e-01 | 3.29e-05 | 1.0 | 1.88e+07 | 9576 | 210 |
| Kenya | 0.33 | 0.4 | 2.74e-01 | 0.e+00 | 1.0 | 5.38e+07 | 1618 | 214 |
| Kuwait | 0.16 | 1.0 | 1.64e-02 | 3.22e-03 | 1.4 | 4.27e+06 | 24112 | 17 |
| Kyrgyzstan | 0.38 | 0.0 | 1.08e-02 | 9.01e-03 | 14.7 | 6.52e+06 | 1594 | -6 |
| Latvia | 0.23 | 1.0 | 6.80e-03 | 1.01e-02 | 133.3 | 1.89e+06 | 1061 | -67 |
| Lebanon | 0.21 | 0.77 | 2.21e-01 | 0.e+00 | 4.8 | 6.83e+06 | 1168 | -92 |
| Lithuania | 0.36 | 1.0 | 4.04e-03 | 2.84e-02 | 230.5 | 2.72e+06 | 1656 | -61 |
| Luxembourg | 0.52 | 0.88 | 2.24e-03 | 0.e+00 | 779.8 | 6.26e+05 | 4008 | -66 |
| Malaysia | 0.27 | 1.0 | 2.29e-04 | 1.45e-02 | 3.0 | 3.24e+07 | 7629 | -62 |
| Maldives | 0.21 | 1.0 | 4.75e-04 | 0.e+00 | 1.0 | 5.41e+05 | 1513 | -22 |
| Mexico | 0.25 | 0.14 | 1.73e-03 | 1.16e-03 | 1000.0 | 1.29e+08 | 81400 | 73 |
| Moldova | 0.5 | 0.47 | 2.25e-03 | 8.16e-03 | 390.8 | 4.03e+06 | 7725 | -29 |
| Morocco | 0.28 | 0.19 | 1.32e-02 | 1.36e-02 | 38.4 | 3.69e+07 | 7643 | -37 |
| Nepal | 0.17 | 0.63 | 1.90e-02 | 0.e+00 | 3.2 | 2.91e+07 | 1042 | 90 |
| Netherlands | 0.41 | 0.7 | 1.14e-03 | 1.78e-02 | 134.5 | 1.71e+07 | 46152 | -57 |
| New Zealand | 1.0 | 0.97 | 2.2e-04 | 7.07e-02 | 110.6 | 4.82e+06 | 1504 | -64 |
| Nigeria | 0.6 | 0.04 | 3.14e-05 | 7.70e-03 | 8.1 | 2.06e+08 | 8915 | -7 |
| Norway | 0.41 | 1.0 | 1.88e-03 | 2.65e-02 | 225.8 | 5.42e+06 | 8411 | -69 |
| Oman | 0.14 | 0.85 | 5.4e-02 | 6.19e-04 | 6.6 | 5.11e+06 | 9009 | 95 |
| Pakistan | 0.2 | 0.29 | 3.7e-03 | 3.88e-03 | 411.0 | 2.21e+08 | 61227 | 10 |
| Panama | 0.46 | 0.35 | 4.60e-03 | 9.15e-03 | 385.2 | 4.31e+06 | 12131 | -36 |
| Peru | 0.33 | 0.18 | 2.02e-03 | 2.06e-03 | 129.1 | 3.3e+07 | 141779 | 5 |
| Philippines | 0.43 | 0.0 | 6.57e-03 | 1.25e-02 | 30.5 | 1.1e+08 | 15588 | -59 |
| Poland | 0.23 | 0.4 | 3.17e-03 | 7.92e-03 | 1000.0 | 3.78e+07 | 22825 | -36 |
| Portugal | 0.98 | 0.04 | 1.82e-03 | 2.22e-02 | 16.6 | 1.02e+07 | 31596 | -58 |
| Qatar | 0.34 | 0.05 | 2.57e-02 | 0.e+00 | 15.6 | 2.88e+06 | 50914 | 50 |
| Romania | 0.44 | 0.39 | 6.52e-04 | 1.33e-02 | 526.3 | 1.92e+07 | 18791 | -46 |
| Russia | 0.23 | 0.7 | 5.41e-04 | 1.42e-03 | 135.5 | 1.46e+08 | 379051 | -21 |
| Saudi Arabia | 0.24 | 0.3 | 3.59e-03 | 3.19e-03 | 154.2 | 3.48e+07 | 80185 | 17 |
| Senegal | 0.29 | 0.72 | 5.71e-05 | 0.e+00 | 500.4 | 1.67e+07 | 3348 | -17 |
| Serbia | 0.46 | 0.23 | 9.88e-03 | 2.47e-02 | 7.4 | 8.74e+06 | 11300 | -49 |
| Singapore | 0.2 | 0.66 | 4.62e-03 | 3.45e-03 | 1.0 | 5.85e+06 | 33249 | -13 |
| Slovakia | 0.41 | 0.44 | 2.58e-01 | 1.24e-02 | 1.0 | 5.46e+06 | 1520 | -54 |
| Slovenia | 1.0 | 0.05 | 7.68e-01 | 5.06e-03 | 1.2 | 2.08e+06 | 1473 | -67 |
| Somalia | 0.68 | 0.23 | 5.58e-01 | 5.61e-03 | 1.2 | 1.59e+07 | 1828 | -25 |
| South Africa | 0.18 | 0.52 | 7.12e-02 | 0.e+00 | 23.5 | 5.93e+07 | 27403 | 127 |
| Spain | 0.69 | 0.65 | 6.76e-01 | 7.94e-03 | 6.7 | 4.68e+07 | 237906 | -60 |
| Sri Lanka | 0.26 | 0.0 | 3.69e-03 | 0.e+00 | 16.1 | 2.14e+07 | 1530 | 350 |
| Sudan | 0.28 | 0.45 | 2.07e-04 | 8.15e-03 | 233.1 | 4.38e+07 | 4346 | 2 |
| Sweden | 0.51 | 0.1 | 2.16e-03 | 7.63e-03 | 311.9 | 1.01e+07 | 35727 | -37 |
| Switzerland | 0.45 | 1.0 | 3.74e-03 | 3.48e-02 | 9.9 | 8.65e+06 | 30796 | -68 |
| Thailand | 0.39 | 0.34 | 2.45e-04 | 1.57e-02 | 1.0 | 6.98e+07 | 3065 | -66 |
| Tunisia | 0.69 | 0.6 | 9.1e-05 | 4.03e-02 | 286.8 | 1.18e+07 | 1068 | -62 |
| Turkey | 0.98 | 0.06 | 1.31e-03 | 2.51e-02 | 202.2 | 8.43e+07 | 160979 | -51 |
| Ukraine | 0.4 | 0.27 | 4.86e-04 | 1.13e-02 | 547.9 | 4.37e+07 | 22382 | -31 |
| UAE | 0.34 | 0.16 | 1.16e-03 | 2.39e-03 | 6.4 | 9.89e+06 | 32532 | -6 |
| United Kingdom | 0.27 | 0.79 | 2.39e-03 | 1.11e-02 | 53.0 | 6.79e+07 | 270508 | -44 |
| US | 0.39 | 0.34 | 3.47e-03 | 8.84e-03 | 1.0 | 3.31e+08 | 1721753 | -45 |
| Uzbekistan | 0.36 | 0.84 | 9.24e-05 | 2.05e-02 | 921.6 | 3.35e+07 | 3444 | -45 |
| Venezuela | 0.14 | 0.86 | 1.18e-04 | 0.e+00 | 706.0 | 2.84e+07 | 1325 | 56 |
| Zambia | 0.37 | 0.39 | 1.84e-04 | 1.31e-02 | 1.0 | 1.84e+07 | 1057 | -10 |
| Guinea-Bissau | 0.34 | 1.0 | 1.51e-03 | 2.61e-02 | 1.0 | 1.97e+06 | 1195 | -24 |
| Mali | 0.15 | 0.96 | 1.19e-02 | 1.22e-02 | 22.7 | 2.03e+07 | 1194 | -25 |
| Tajikistan | 0.24 | 1.0 | 6.80e-02 | 2.66e-02 | 18.7 | 9.54e+06 | 3563 | -9 |

**Figure 4:**
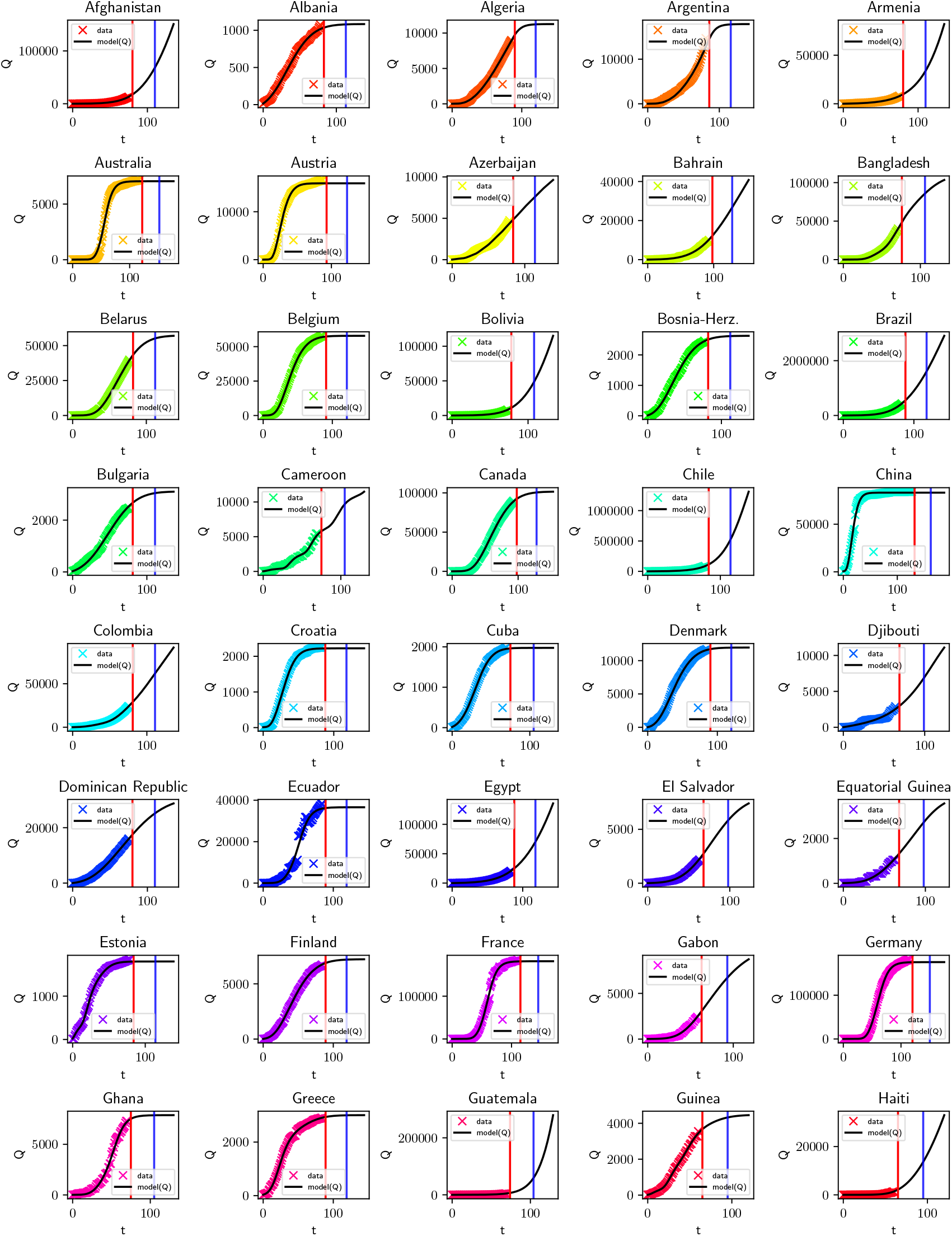
Number of confirmed cases (coloured markers) vs Model (Black line) for countries in alphabetical order from Afghanistan to Haiti. The red vertical line corresponds to 1st June 2020 and the blue vertical line correspond to 1st August 2020.

**Figure 5:**
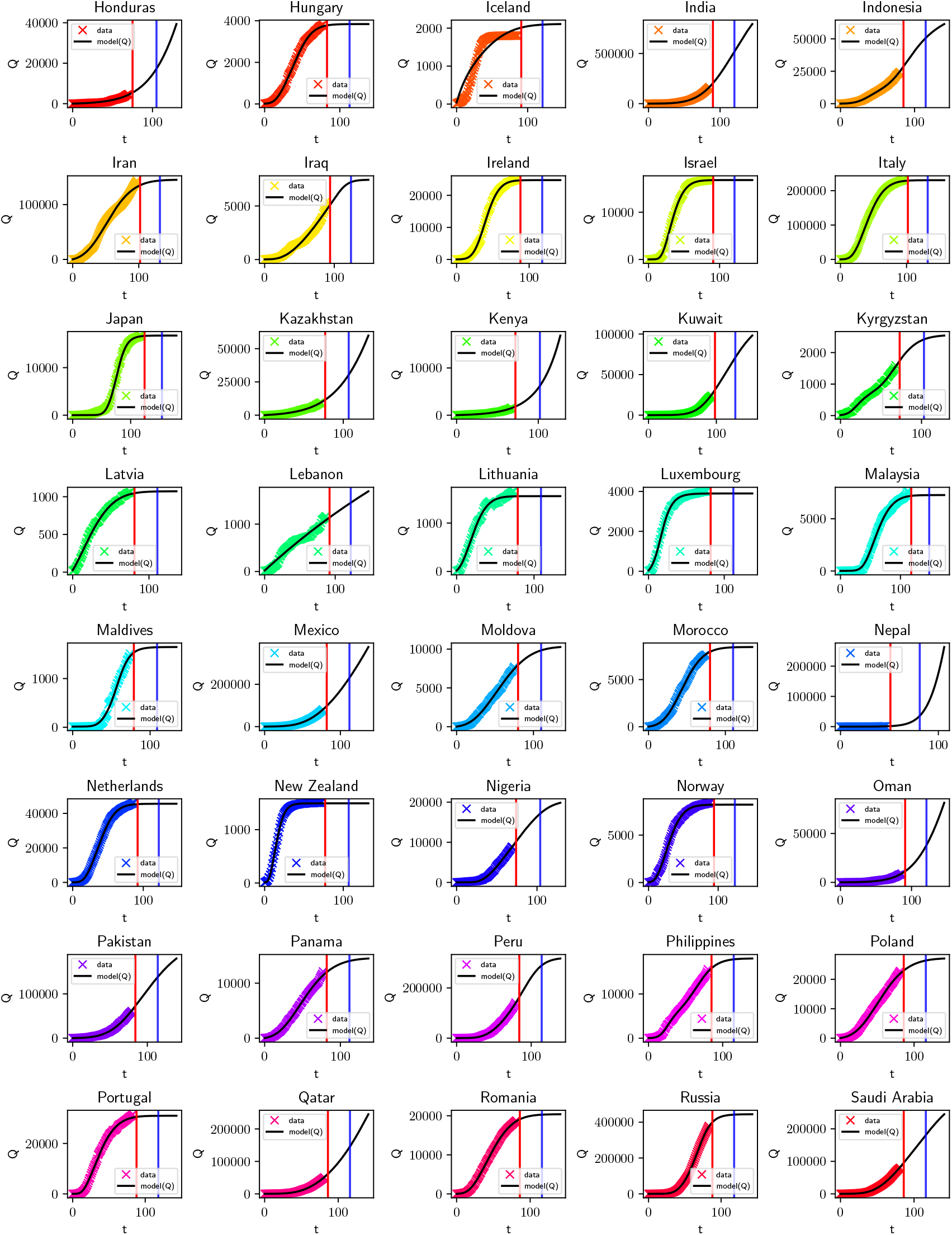
Number of confirmed cases (colored markers) vs Model (Black line) for countries in alphabetical order from Honduras to Saudi Arabia. The red vertical line corresponds to 1st June 2020 and the blue vertical line correspond to 1st August 2020.

**Figure 6:**
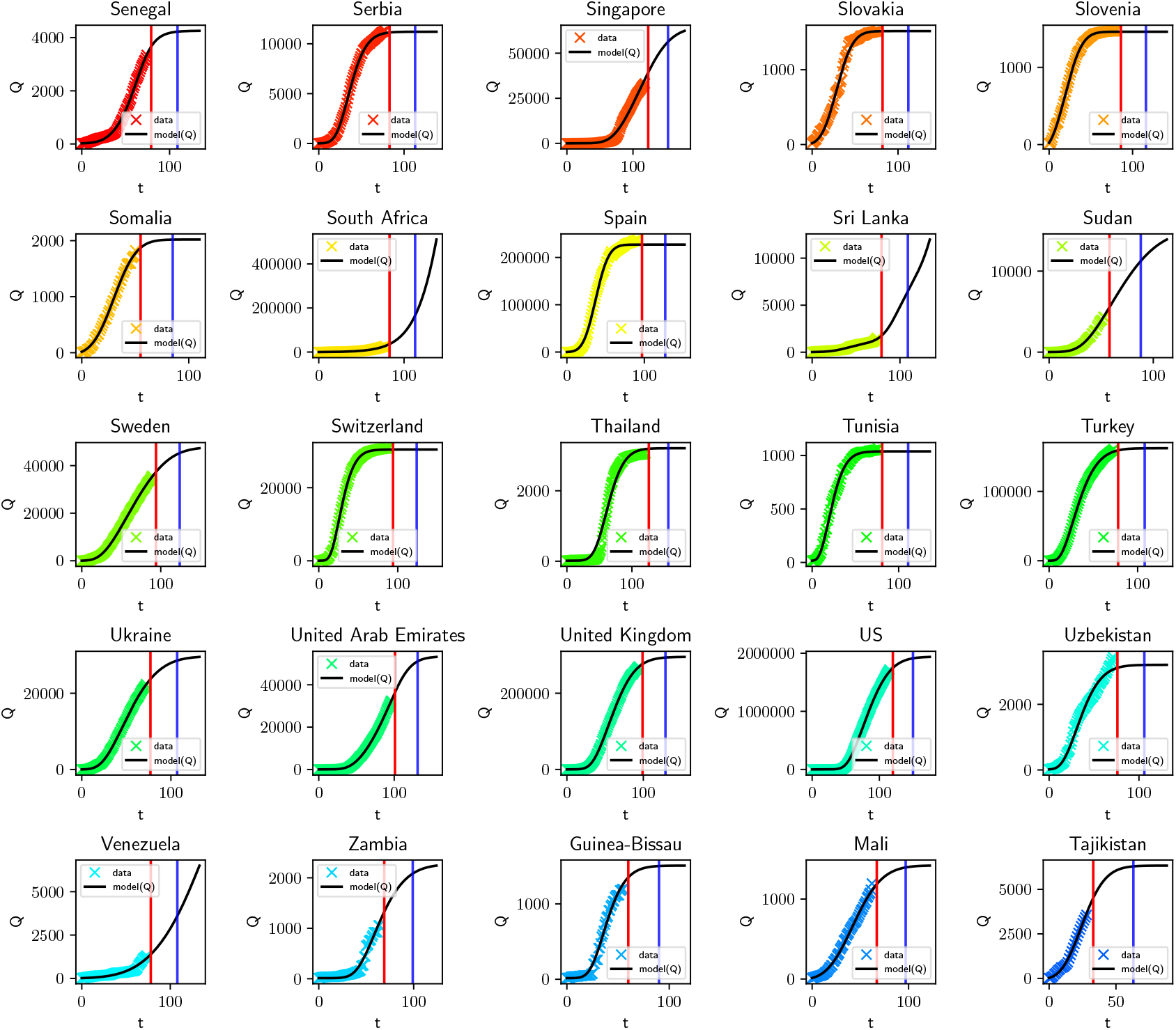
Number of confirmed cases (colored markers) vs Model (Black line) for countries in alphabetical order from Senegal to Tajikistan. The red vertical line corresponds to 1st June 2020 and the blue vertical line correspond to 1st August 2020.

#### 3.1.1 List of Country-Specific model parameters

Table 2 lists the model parameters for over 105 countries. Using the model parameters, we used Eqs.(1–5) to solve for the maximum number of infections, the corresponding number of days from 1st June 2020 was identified and is listed as *t_maxI_* in the table 2.

According to the proposed model, the parameter *β* is proportional to transmission and contact activities. *μ^0^→* 0 corresponds to low inter-cluster transmission and *μ^0^→* 1 corresponds to high inter-cluster transmission. If *ρ < κ* the primary mitigation is using lockdown restrictions.

If *ϕ*_0_ *→* 1 the number of untested active infections is low while *ϕ*_0_ *→* 1000 corresponds to a high-number of untested active infections. However, due to the interactions and correlations among the parameters, we would not recommend interpreting the actual efficiency of the mitigation measure of a country based on these parameters. Estimates of these parameters can be improved with additional data. Also listed in the table is the total number of reported confirmed cases 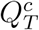 that serves as an indicator for the volume of data used in the fitting procedure.

### 3.2 Lockdown continuation

The list of countries that should NOT relax lockdown restrictions is provided in Table 3. This list is based on the model prognosis of the date of maximum number of active infections that lies in the future as of 1 June 2020. These countries should definitely continue and tighten lockdown restrictions until after the predicted peak date. See the next section for a qualitative analysis of the role of lockdown duration and relaxation rate on a cluster-specific evolution of the pandemic.

**Table 3:**
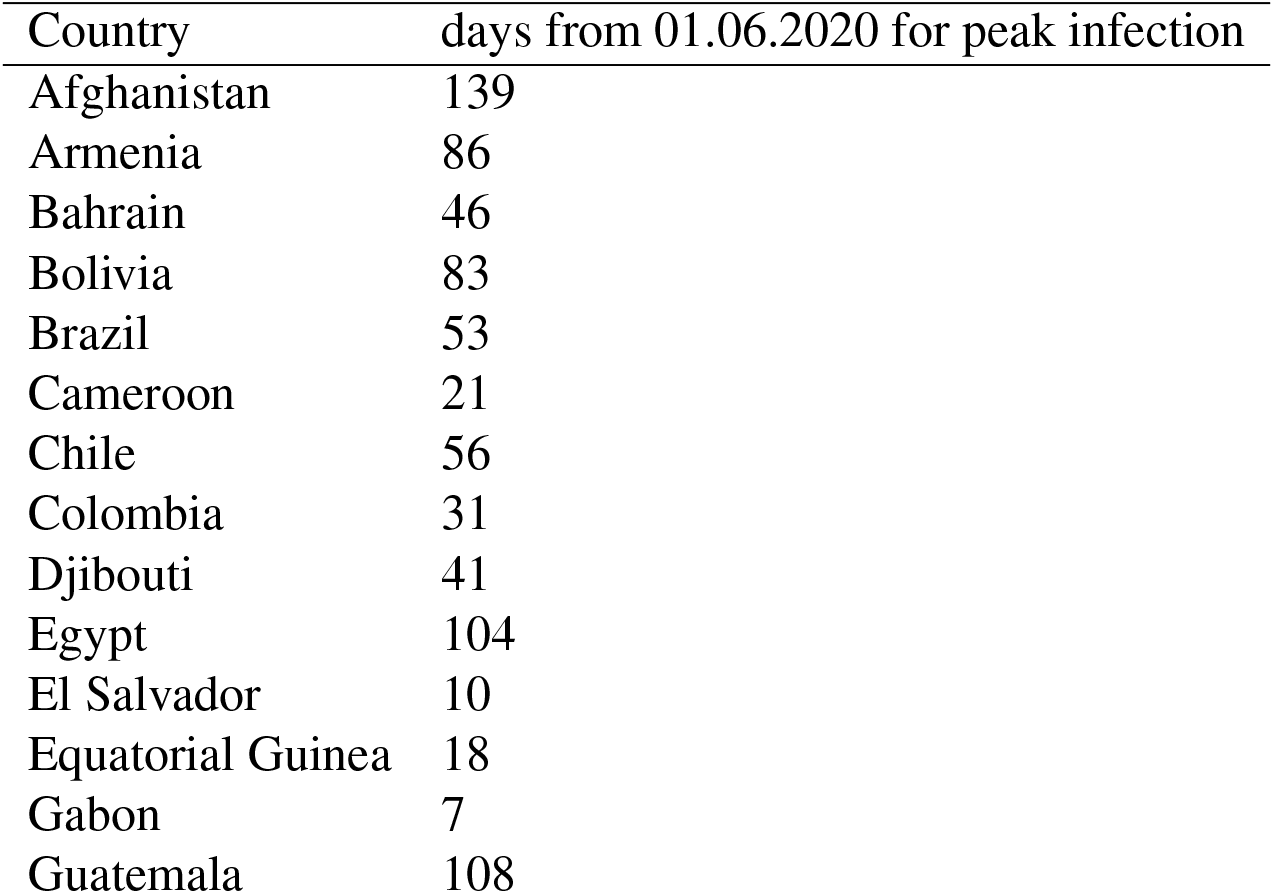
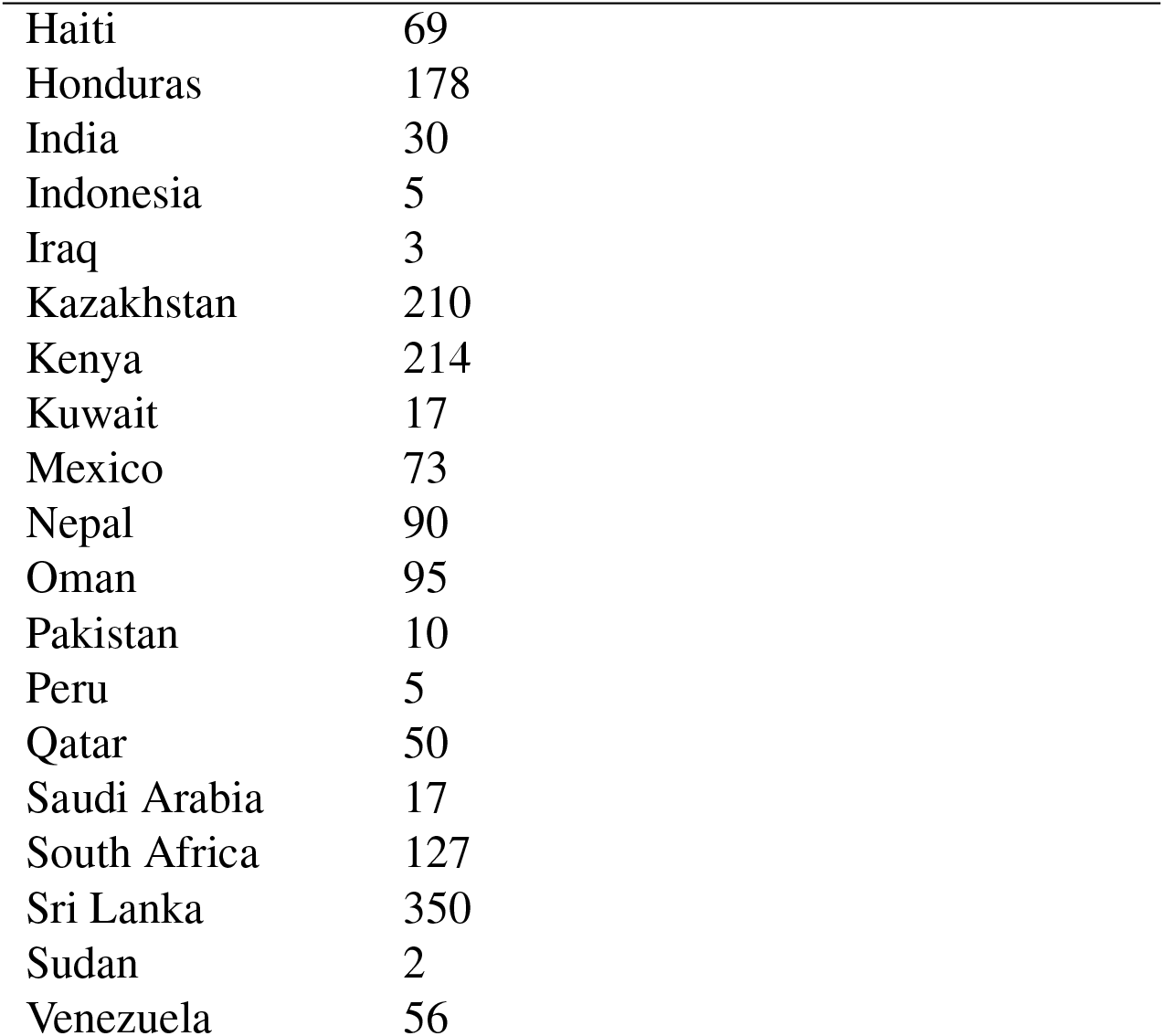
List of countries that should NOT relax lockdown restrictions: Model-based projections

| Country | days from 01.06.2020 for peak infection |
| --- | --- |
| Afghanistan | 139 |
| Armenia | 86 |
| Bahrain | 46 |
| Bolivia | 83 |
| Brazil | 53 |
| Cameroon | 21 |
| Chile | 56 |
| Colombia | 31 |
| Djibouti | 41 |
| Egypt | 104 |
| El Salvador | 10 |
| Equatorial Guinea | 18 |
| Gabon | 7 |
| Guatemala | 108 |
| Haiti | 69 |
| Honduras | 178 |
| India | 30 |
| Indonesia | 5 |
| Iraq | 3 |
| Kazakhstan | 210 |
| Kenya | 214 |
| Kuwait | 17 |
| Mexico | 73 |
| Nepal | 90 |
| Oman | 95 |
| Pakistan | 10 |
| Peru | 5 |
| Qatar | 50 |
| Saudi Arabia | 17 |
| South Africa | 127 |
| Sri Lanka | 350 |
| Sudan | 2 |
| Venezuela | 56 |

### 3.3 Lockdown relaxation

The list of countries whose date of peak active infections lies before 1 May 2020 is listed in Table 4. However, even after the date of maximum number of active infections, the rate of decline of the number of active infections is not the same and varies from country to country. Thus, it is recommended that the countries relax lockdown restrictions in proportion to the rate of decline of active infections shown in the figures 12, 13 and 14. See the next section for a qualitative analysis of the role of lockdown duration and relaxation rate on the cluster-specific evolution of the pandemic.

**Table 4:**
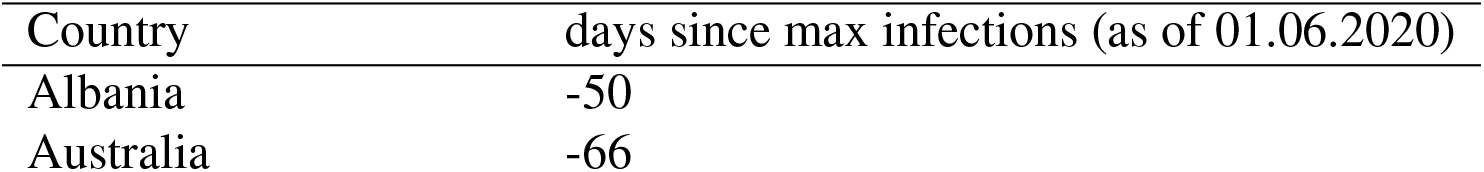
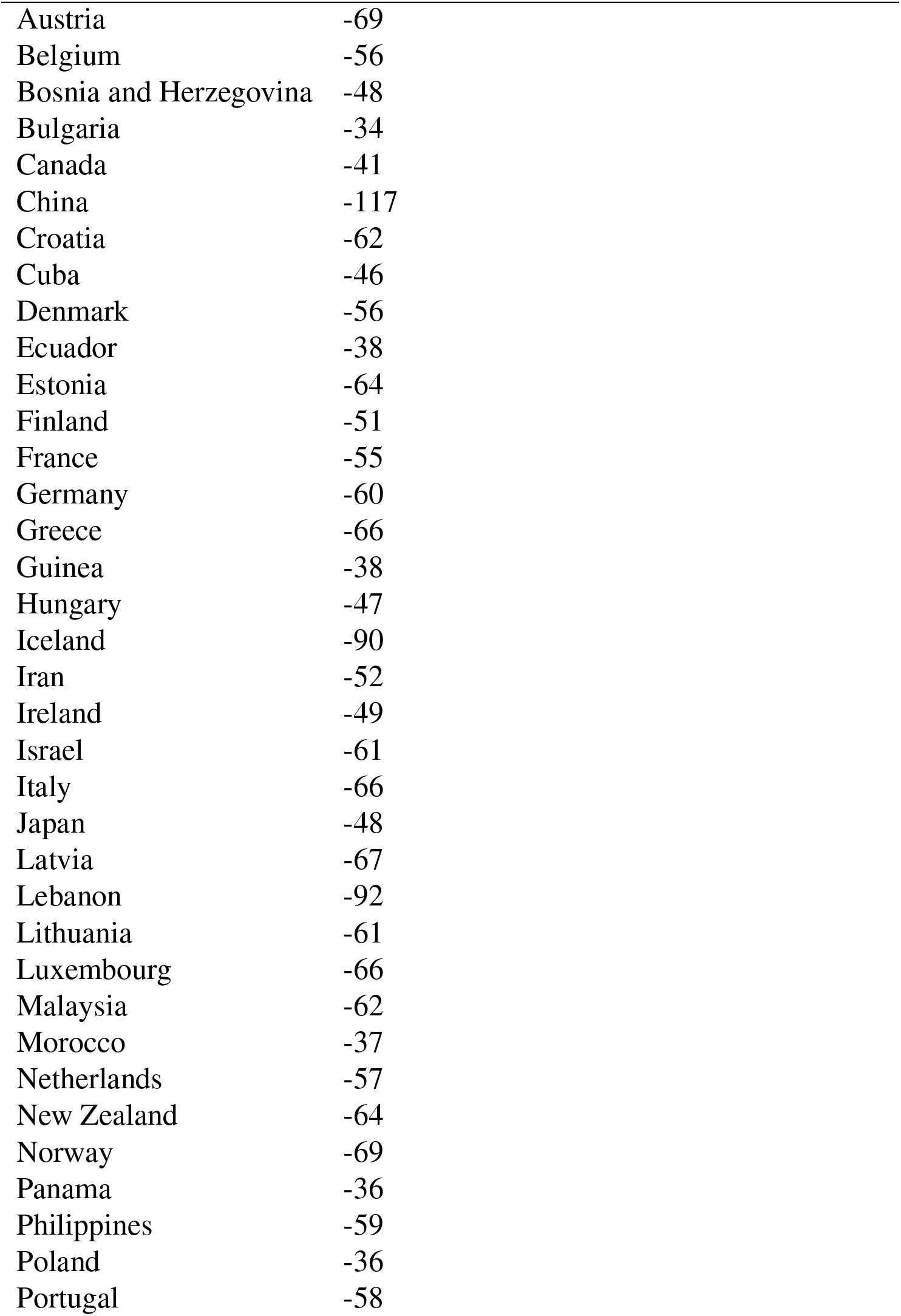
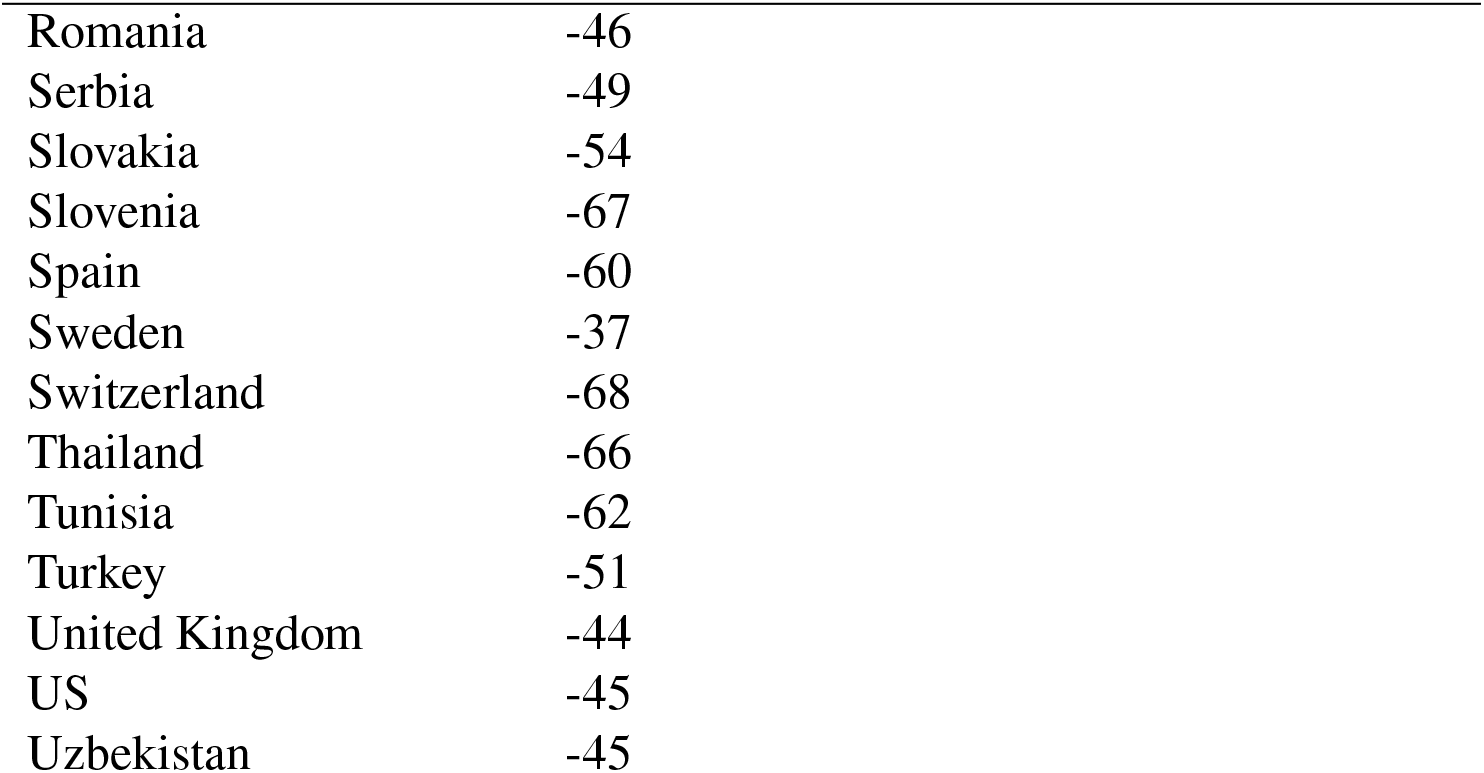
List of countries that can relax lockdown and other restrictions proportional to the predicted rate of decline of active infections: Model based prognosis

| Country | days since max infections (as of 01.06.2020) |
| --- | --- |
| Albania | -50 |
| Australia | -66 |
| Austria | -69 |
| Belgium | -56 |
| Bosnia and Herzegovina | -48 |
| Bulgaria | -34 |
| Canada | -41 |
| China | -117 |
| Croatia | -62 |
| Cuba | -46 |
| Denmark | -56 |
| Ecuador | -38 |
| Estonia | -64 |
| Finland | -51 |
| France | -55 |
| Germany | -60 |
| Greece | -66 |
| Guinea | -38 |
| Hungary | -47 |
| Iceland | -90 |
| Iran | -52 |
| Ireland | -49 |
| Israel | -61 |
| Italy | -66 |
| Japan | -48 |
| Latvia | -67 |
| Lebanon | -92 |
| Lithuania | -61 |
| Luxembourg | -66 |
| Malaysia | -62 |
| Morocco | -37 |
| Netherlands | -57 |
| New Zealand | -64 |
| Norway | -69 |
| Panama | -36 |
| Philippines | -59 |
| Poland | -36 |
| Portugal | -58 |
| Romania | -46 |
| Serbia | -49 |
| Slovakia | -54 |
| Slovenia | -67 |
| Spain | -60 |
| Sweden | -37 |
| Switzerland | -68 |
| Thailand | -66 |
| Tunisia | -62 |
| Turkey | -51 |
| United Kingdom | -44 |
| US | -45 |
| Uzbekistan | -45 |

## 4 Individual-Based Simulation of Lockdown

A realistic description of spatio-temporal dynamics (i.e movements) of individuals and their interactions in a typical cluster i.e. city, town or village is quite complex. However, an idealized generic manner of interaction of individuals consists of a) regular interaction with family members, b) interaction with individuals at the work/study place, c) interaction during travel to work/study and d) interaction during shopping. Thus, the individual-based model consists of 4 layers, family layer, commuting layer (transport), workplace/school layer and public place layer where individual-individual interactions take place. The virtual population is constructed by specifying the total number of families and a family-size distribution, a transport size distribution and a work/study place distribution. A family-size distribution allows to model families of different sizes, the transport size distribution allows for modelling different types of commuting systems (taxi, car, train, bus) and the work/study place distribution allows to model firms, small-businesses, offices, schools and universities (see Fig. 11).

### 4.0.1 Individual dynamics

Mobility of individuals is realized by creating a simulation day. Each simulation day consists of the following stages. Individuals first move from the family layer to the transport layer (not all members of the family are mobile. This is specified using an additional parameter). Thereafter, the individuals move from the transport layer to the work/study place layer, then they return to the family layer using the same sequence and then move to the public-place layer and then back to the family layer. The different layers are depicted in Fig. 7.

**Figure 7:**
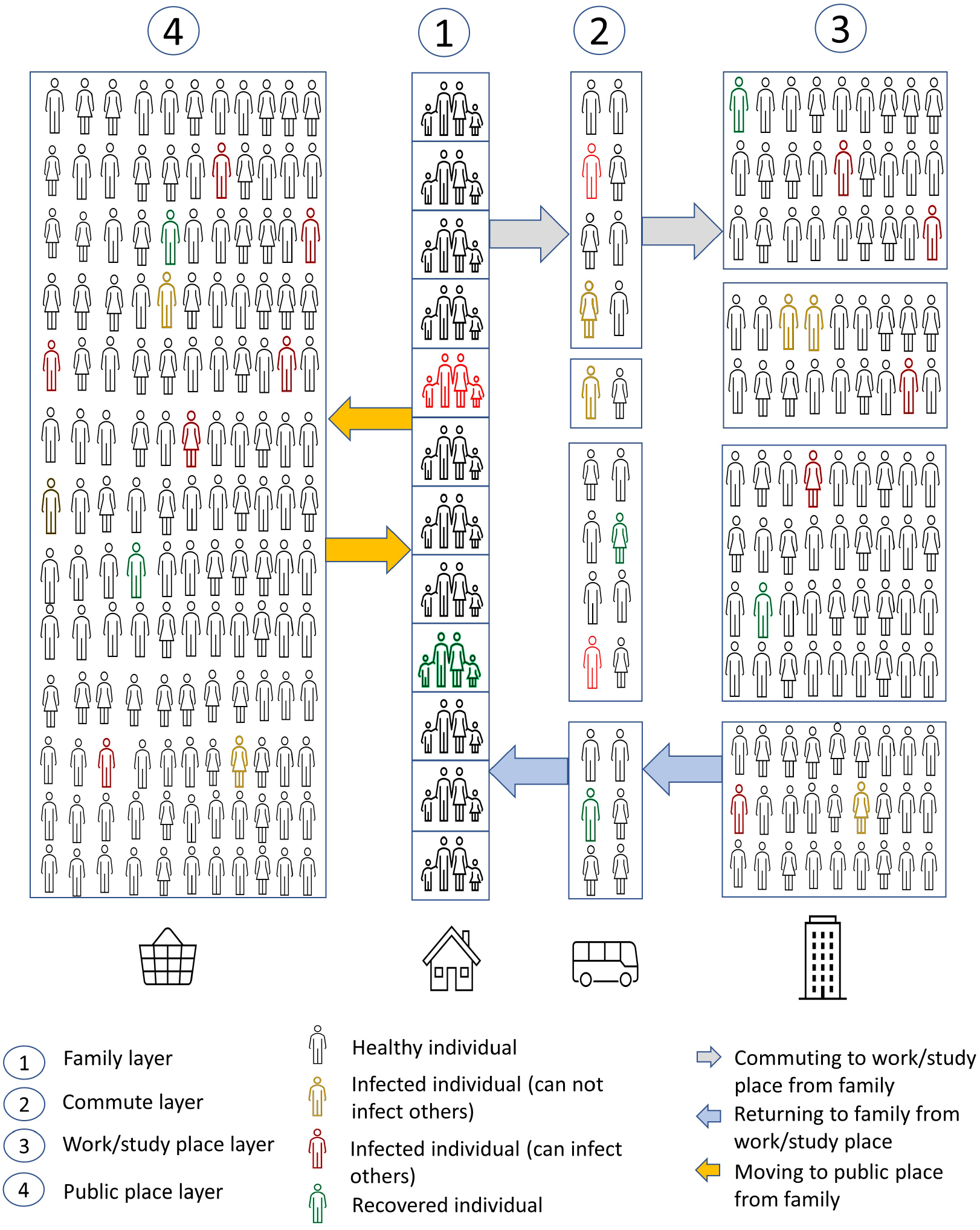
Schematic of the Individual-Based model for disease spreading in communities

### 4.0.2 Disease dynamics

In each layer an infection sweep is performed to check for possible infection. A healthy individual gets infected through contact with an infected individual based on a certain parameter *p* which is the probability of infection. Once infected, each individual goes through the following stages of disease progression from the first day of infection. The individual once exposed, after time *t_i_* is infectious. These individuals remain infectious for a time period *t_I_* and recover after a time period *t_R_*. Symptomatic individuals die after a time period *t_F_*. The range of values used in the simulation are provided in table 5. We assume that 50 percent of the infected population are asymptomatic (13). We also modify this fraction of asymptomatic individuals in the final analysis.

**Table 5:** List of Model Parameters

| Description | Value |
| --- | --- |
| Population size | 1million |
| Probability that a symptomatic individual dies (CFR) | 6 % |
| Time to infectiousness | 2 to 4 days |
| Incubation time | 3 to 14 days (4) |
| Time to recover for asymptomatic patients | 7 to 21 days |
| Time to fatality or recovery for symptomatic patients | 8 - 60 days (4) |

### 4.0.3 Interactions

A family is characterized by number of members per family and number of mobile members who go out to work/study place. One person from each family goes to public place on every simulation day. If an individual in a certain family is infected, all the members of the family are infected. Once the infected individuals show symptoms, they automatically self-quarantine. In order to simulate interaction during travel to work/study, depending on the commuting service assigned to the individual, the individual is placed randomly in a rectangular grid of cells whose size is proportional to the size of the transport vehicle. Each individual has a fixed mode of transportation on each simulation day but his position in the transport vehicle changes everyday. Once placed in this rectangular grid, an infected individual infects individuals in the cells that belong to the Moore neighbourhood of the cell associated with cell of the infected individual with probability of infection *p*. This means we generate a random number for each potential infection and if this random number is smaller than the *p*, the individual is infected. In order to simulate interaction at the work/study place, the individual is placed on a cell in a rectangular grid of cells that represent a unique work/place associated with the specific individual. This grid of cells is unique for each individual. The size of the grid varies based on the number of employers per work place or students per school/university. The manner of interaction of infected and healthy individuals here is similar to that during transport with same probability of infection *p*. In contrast to the situation during transportation where the cell is not associated with the individual every day, during work/study, the cell associated with a particular individual is permanent. i.e the individual is placed in the same cell every day until quarantined or dead. Finally interaction in the public layer is as follows. Given the total number of individuals who will visit the public layer, a single rectangular grid similar to work/study place is created. The individual is placed randomly in a cell in this grid. Here the mechanism of infection is same as in the transport layer and the work/study place layer.

### 4.0.4 Numerical Experiments

#### Piece-wise Linear Lockdown

To simulate the influence of lockdown in a population that strictly adheres to the government mandated lockdown regulations, we assume the probability of infection *p* to be a piece-wise linear function of time. Due to social distancing restrictions (and other factors such as hand-washing etc.), the probability that an individual is infected also decreases. Thus we simulate this lockdown as the change in probability of infection that starts at *p* = 1, decreases to *p* = *p_min_*, stays at *p_min_* for a specified duration and then goes back to *p* = 1. Fig. 8 shows the various levels of population normalized infection for various piece-wise linear lockdown scenarios. The background colors in each plot in Fig. 8 characterizes the lockdown duration and level. The purple region corresponds to the decrease stage *p* to *p_min_*. The green region corresponds to constant *p_min_*. The light red region correspond to the release of lockdown from *p_min_* to *p* = 1. Fading green corresponds to *p_min_* = 0.001 and bright green corresponds to *p_min_* = 0.0001. We assume that the duration from *p → p_min_* is one week. We consider three different lockdown durations, 21 days, 49 days and 77 days and three different lockdown release rates of 0.1, 0.01 and 0.001. These 9 simulations we run for two different lockdown intensities i.e. two different *p_min_*. Thus we investigate lockdown intensity, duration and release rate. In general as expected more the intensity and longer the lockdown duration, more effective is the procedure in mitigating the spread of the disease. However the most interesting effect is the influence of the release rate. According to plots a,b,c and j,k,l a 21 day lockdown is inefficient in reducing the total number of active infections irrespective of the lockdown intensity. However we see a delay in the peak of maximum infections as seen in plots c and l. Thus the absence of new infections when a certain lockdown procedure is implemented does not guarantee that the infection is completely eliminated. It could be the case that new infections can emerge and exponentially grow once the lockdown is completely lifted. This delay also depends on the release rate. For much smaller release rates, we do not expect to see this effect.

**Figure 8:**
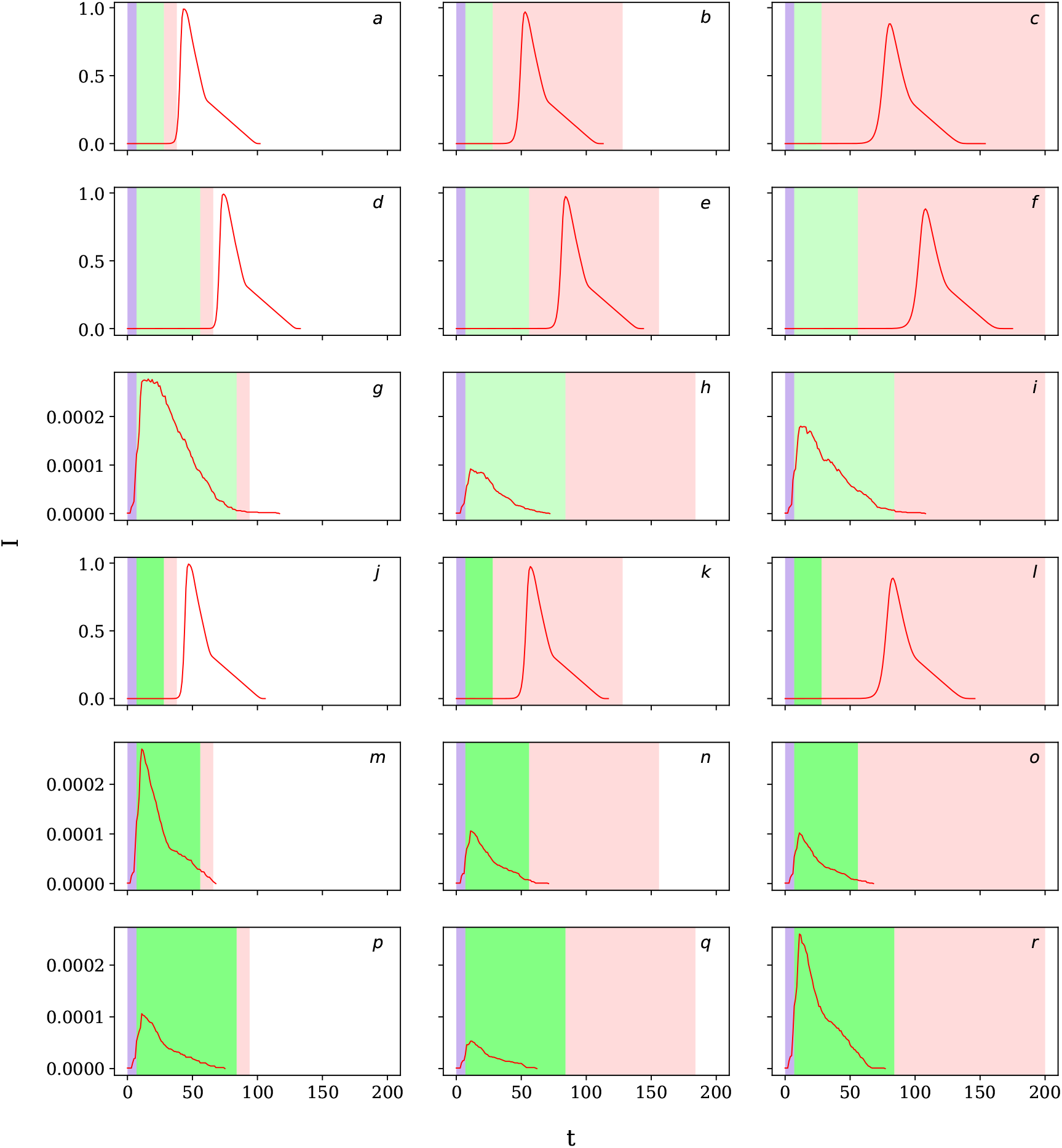
Population normalized infection for various types of piece-wise linear lockdown procedures as a function of time. The background colors characterize the lockdown duration and level. The purple region corresponds to the decrease stage *p* to *p_min_*. The green region corresponds to constant *p_min_*. The light red region correspond to the release of lockdown from *p_min_* to *p* = 1. Fading green corresponds to *p_min_* = 0.001 and bright green corresponds to *p_min_* = 0.0001.

#### Exponential-Log lockdown

While the aforementioned lockdown curves are assumed to be piece-wise linear and are assumed to represent a population that strictly adheres to the lockdown policy of the government, in certain cases, the population attempts to adhere to the policies but would remain in a strict state of lockdown only for a very short period. We model this situation using an exponential-log lockdown curve according to the following equation:

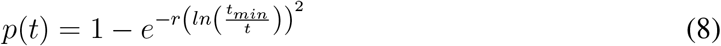

In this equation *t_min_* is the time when *p* = *p_min_* and *r* is the lockdown release rate constant. Using these parameters we can model a variety of lockdown scenarios. Fig. 9 shows the spread of the disease as a function of time for *t_min_* = 21 days (left), *t_min_* = 42 days (center) and *t_min_* = 56 days (right) for four different values of *r* as 1, 0.1, 0.05, 0.05. In contrast to piecewise lockdown, in case of exponential-log lockdown, there is only a temporal shift in the disease and no apparent reduction in the number of active infections is observed. This is in part due to the short-lived *p_min_*.

**Figure 9:**
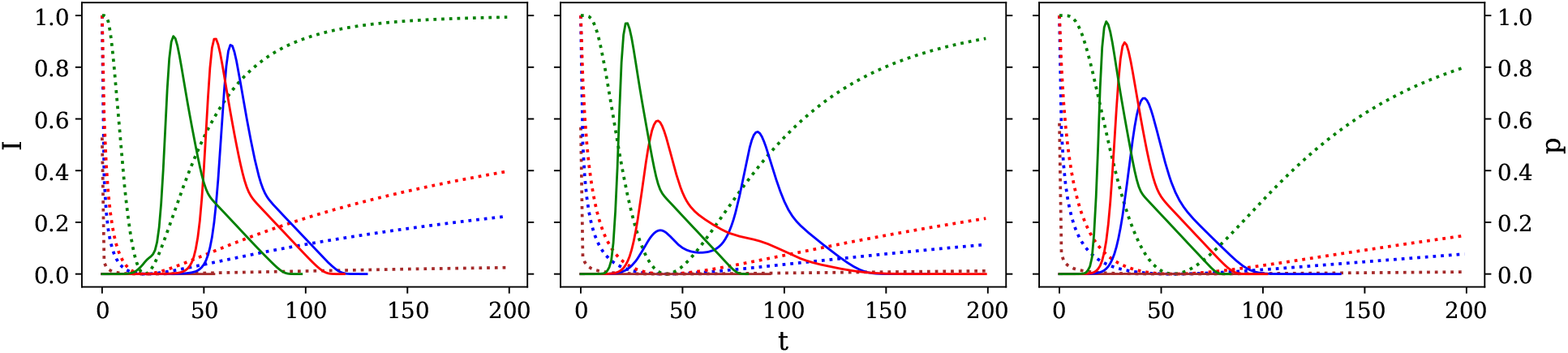
Population normalized infection for various types of Exponential-log lockdown procedures as a function of time.

#### Influence of Asymptomatic population

In order to obtain insight into the role of the hidden asymptomatic infectious individual we vary the proportion of asymptomatic individuals in the population. Fig. 10 shows the effect of the level of asymptomatic infected populations on the overall level of active infections in conjunction with various piece-wise lockdown procedures assuming *p_min_* = 0.001. The analysis shows that a similar behaviour is to be expected for an asymptomatic proportion of 25 percent or 50 percent.

**Figure 10:**
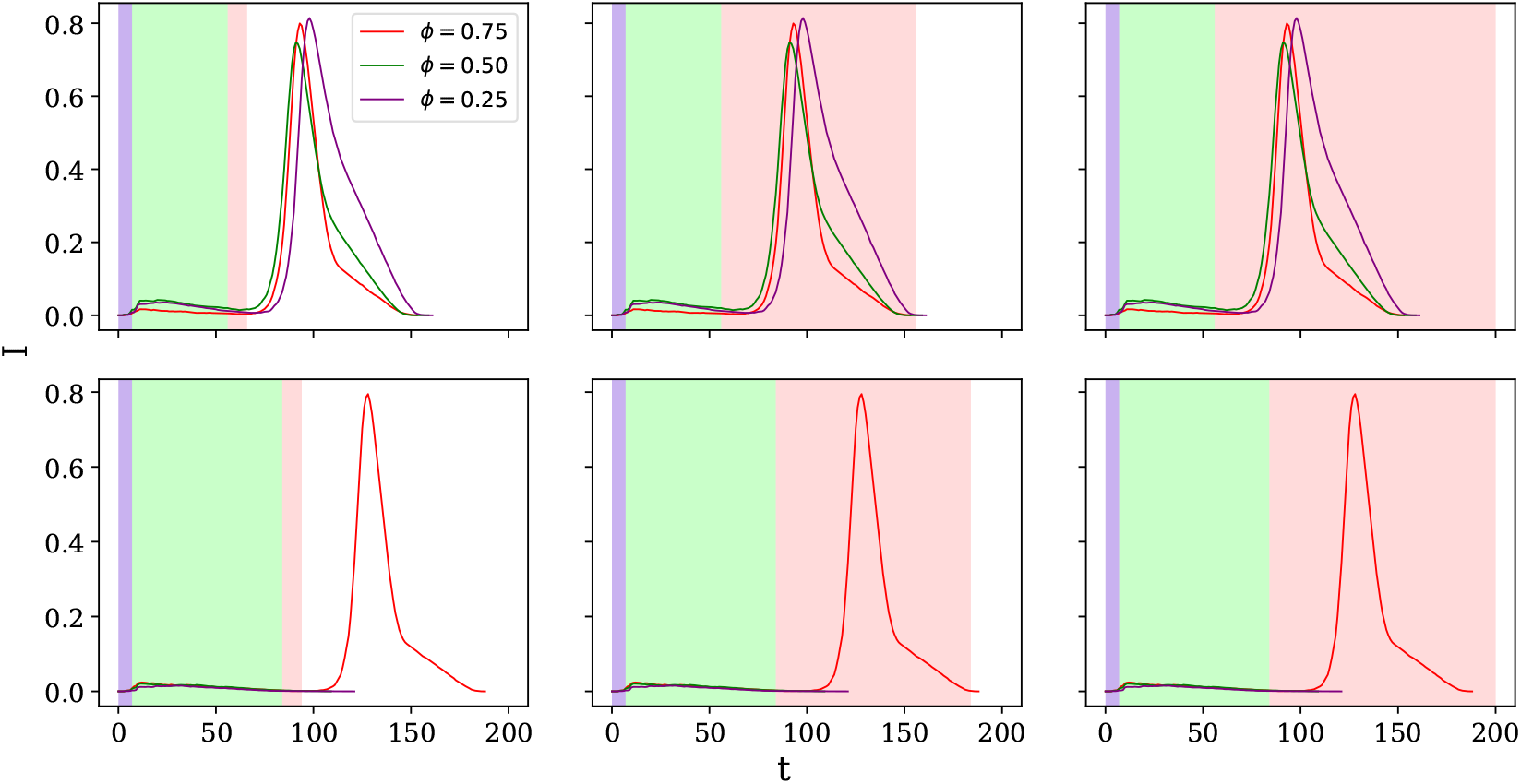
Population normalized infection for various types of piece-wise lockdown procedures and various levels of asymptomatic populations as a function of time.

## 5 Conclusion

We have investigated the spread of COVID-19 in small and large populations. Based on our analysis we come to the following conclusions.

1. The proposed Lattice-SIRQL model is a hybrid formulation suitable for large-scale analysis with a certain degree of spatial interactions.
2. A wide range of microscopic effects can manifest a similar behaviour at the macroscopic (global) scale especially when mitigation measures are considered.
3. We calibrate the Lattice-SIRQL model for over 105 countries to make country-specific recommendations for lockdown continuation or lockdown relaxation. We also provide indicators for possible evolution of active infections based on model prognosis.
4. In addition to the phenomenological Lattice-SIRQL model we also simulate the spreading of the disease using a microscopic, cluster-level, individual-based model that takes into account the complex dynamics of a population. As each individual is modelled explicitly, the simulation is limited to small populations < 1 million. Numerical experiments on the mechanics of lockdown effects have been investigated.
5. The results presented here are only guidelines that provide one of the different ways COVID-19 could evolve. With regards to the reliability of our model projections we quote Dr. Fauci (7) *”you don’t make the timeline, the virus makes the timeline”*.

## Data Availability

Computer code used in the paper is available on request.

## Appendix

### Population distribution for the Individual-based model

### Model Prognoses for active infectious cases

The model prognosis using the identified parameters are provided in figures 12, 13 and 14. While predictions for most countries are within the realistic range of the expected behaviour, the predictions for the following countries are suspect. Cameroon, Lebanon and Sri Lanka. The non-monotonous variations in the case of Cameroon, the unrealistic reduction in cases in Lebanon and the extremely large increase in the cases in Sri Lanka are in the authors opinion not as expected. As already mentioned the model heavily relies on the number of reported cases, secondly more data could help provide an improved correct estimation for these countries.

**Figure 11:**
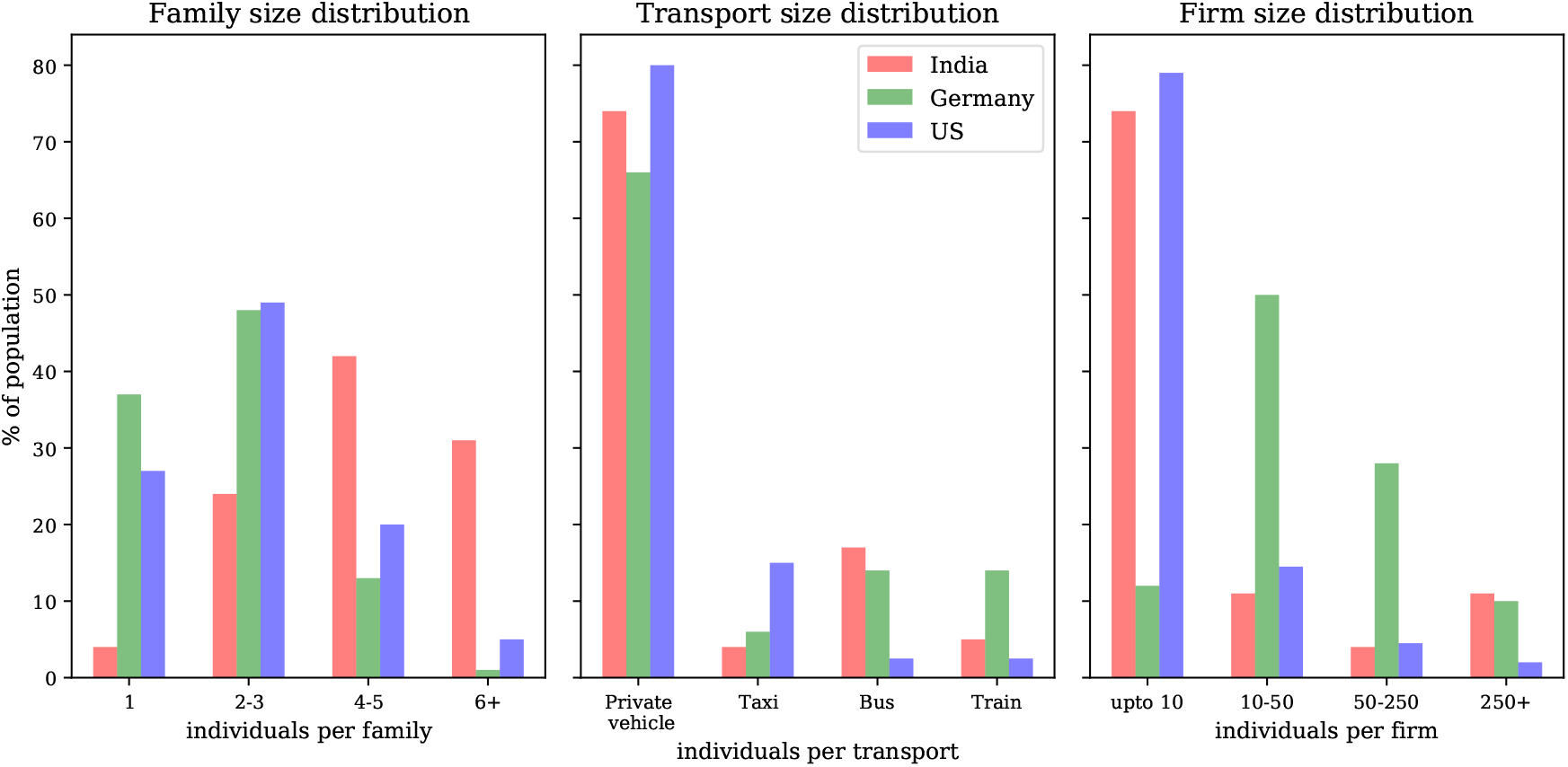
Distribution of transport systems in Germany, India and USA

**Figure 12:**
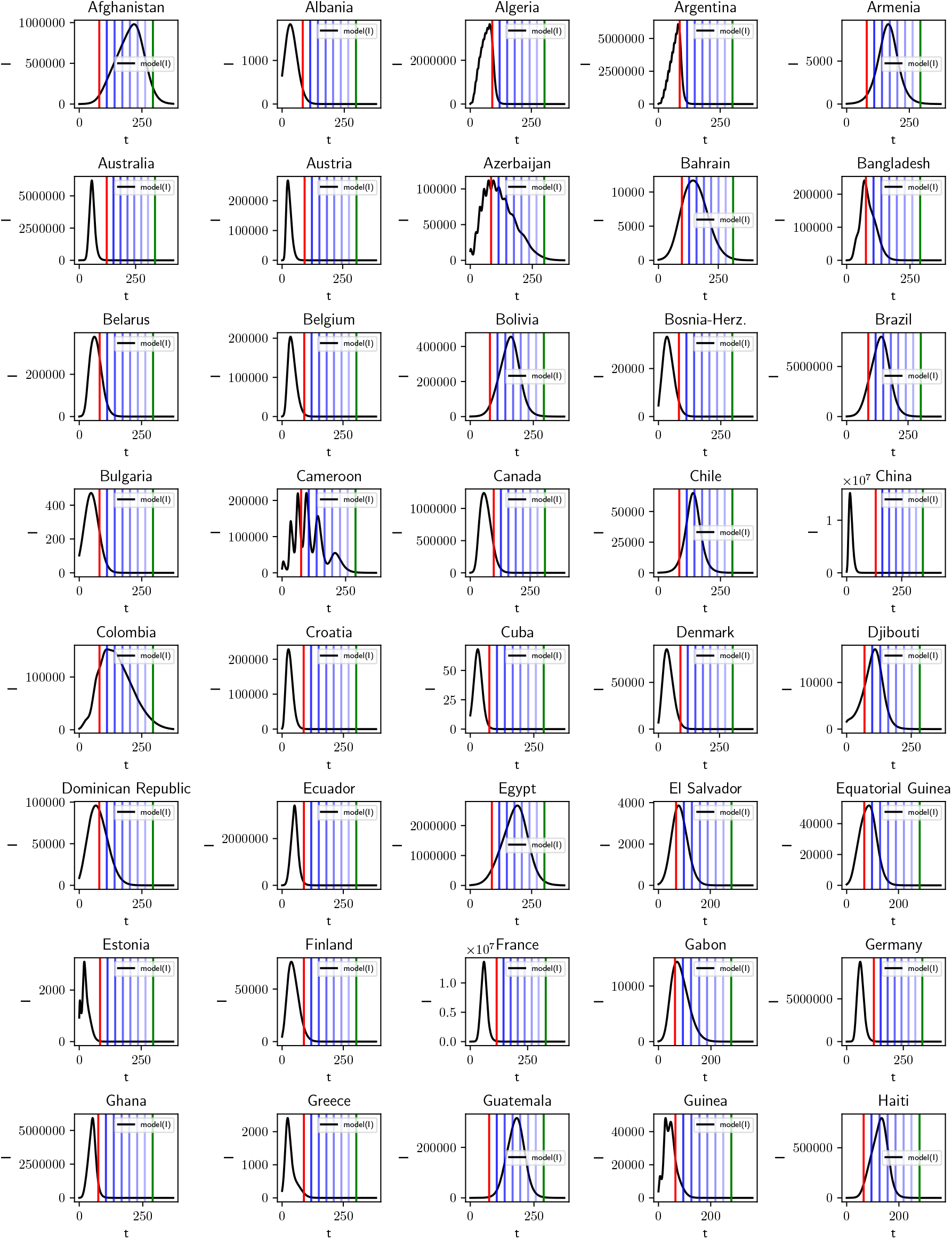
Model predictions of the number of active COVID-19 cases from 1st June 2020 (red vertical line) until 31st December 2020 (green vertical line) and beyond. Predictions for Cameroon has to be interpreted with caution.

**Figure 13:**
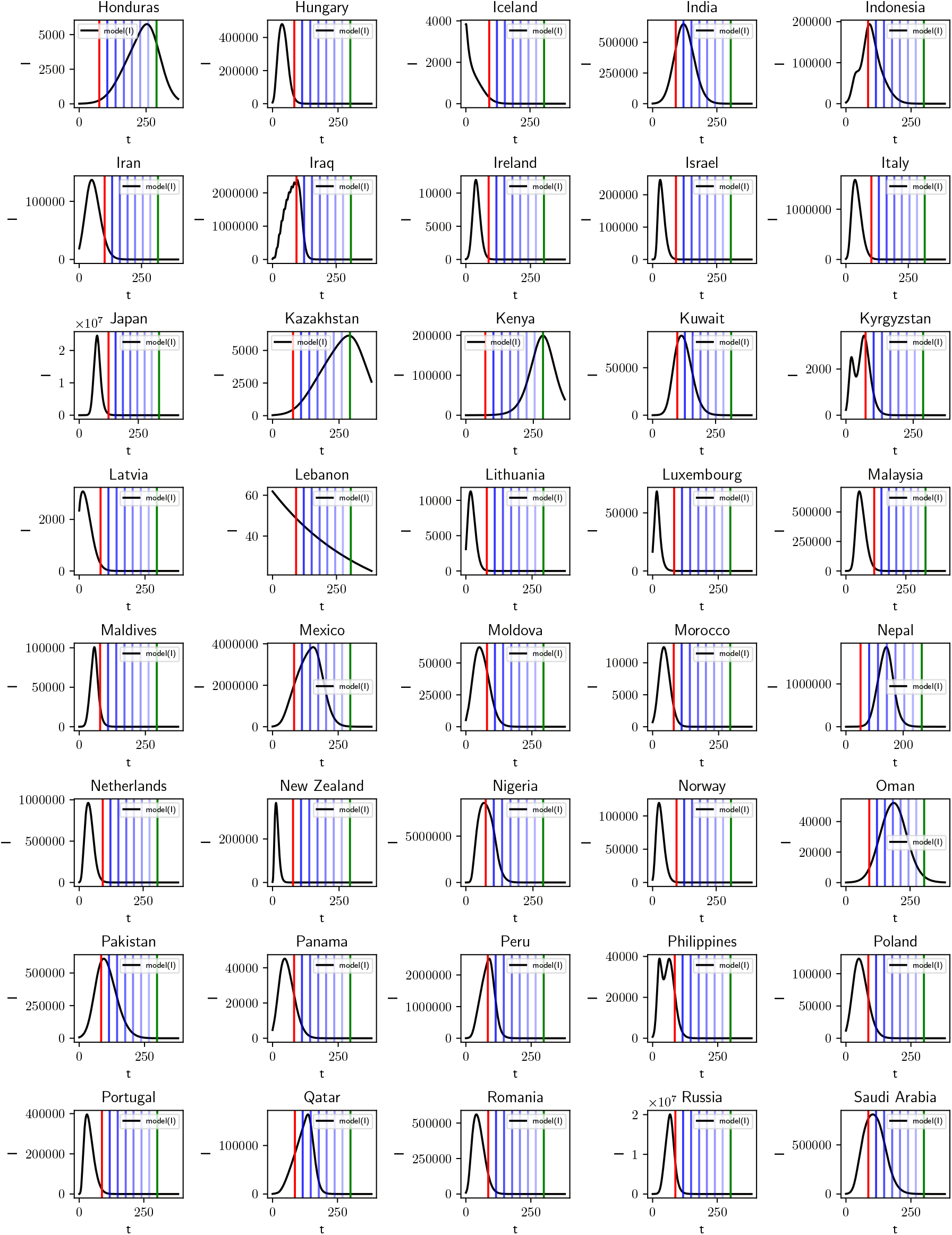
Model predictions of the number of active COVID-19 cases from 1st June 2020 (red vertical line) until 31st December 2020 (green vertical line) and beyond. Predictions for Lebanon is to be discarded.

**Figure 14:**
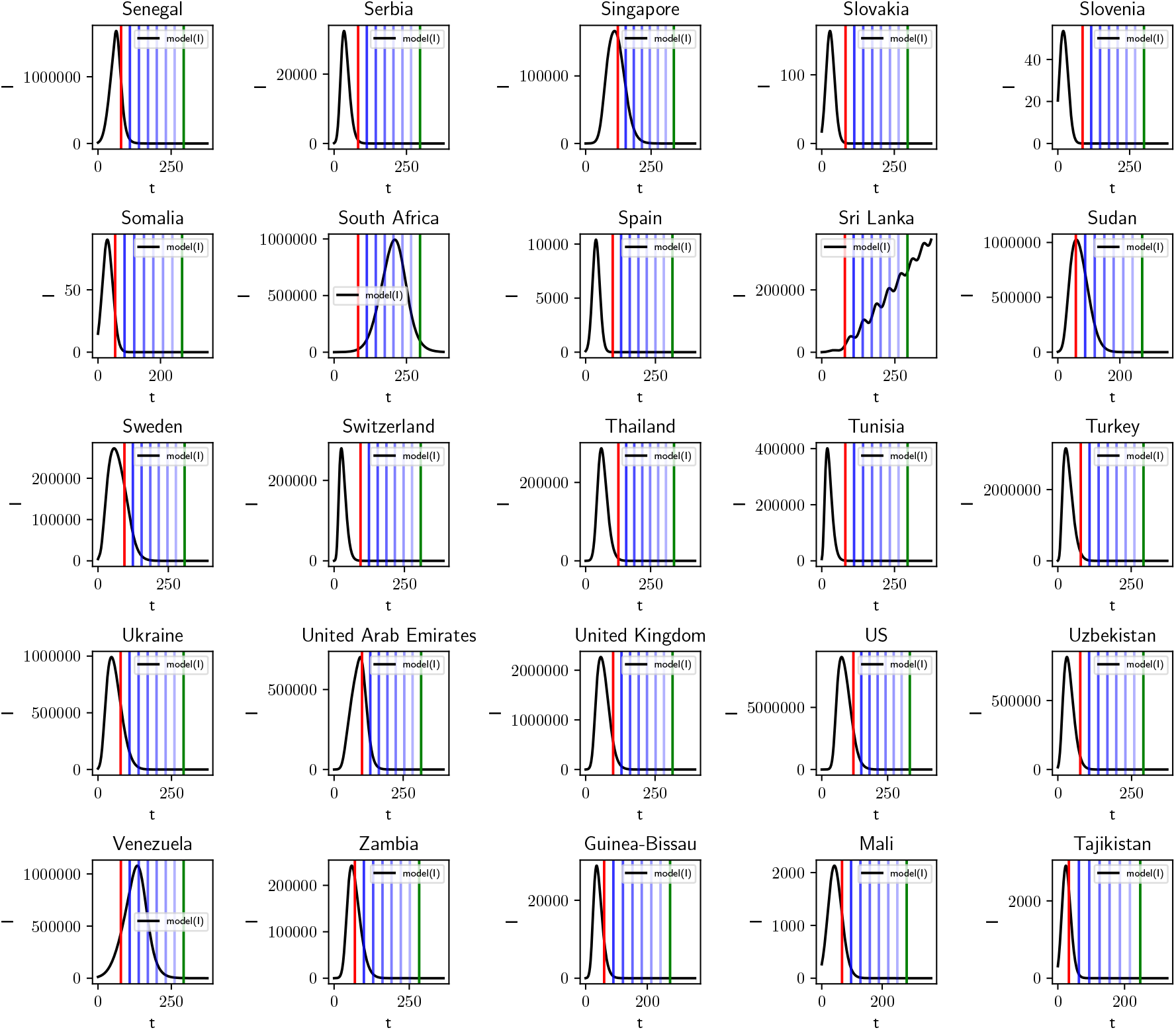
Model predictions of the number of active COVID-19 cases from 1st June 2020 (red vertical line) until 31st December 2020 (green vertical line) and beyond. Predictions for Sri Lanka is to be interpreted with caution.

